# Researching COVID to enhance recovery (RECOVER) pediatric study protocol: Rationale, objectives and design

**DOI:** 10.1101/2023.04.27.23289228

**Authors:** Rachel Gross, Tanayott Thaweethai, Erika B. Rosenzweig, James Chan, Lori B. Chibnik, Mine S. Cicek, Amy J. Elliott, Valerie J. Flaherman, Andrea S. Foulkes, Margot Gage Witvliet, Richard Gallagher, Maria Laura Gennaro, Terry L. Jernigan, Elizabeth W. Karlson, Stuart D. Katz, Patricia A. Kinser, Lawrence C. Kleinman, Michelle F. Lamendola-Essel, Joshua D. Milner, Sindhu Mohandas, Praveen C. Mudumbi, Jane W. Newburger, Kyung E. Rhee, Amy L. Salisbury, Jessica N. Snowden, Cheryl R. Stein, Melissa S. Stockwell, Kelan G. Tantisira, Moriah E. Thomason, Dongngan T. Truong, David Warburton, John C. Wood, Shifa Ahmed, Almary Akerlundh, Akram N. Alshawabkeh, Brett R. Anderson, Judy L. Aschner, Andrew M. Atz, Robin L. Aupperle, Fiona C. Baker, Venkataraman Balaraman, Dithi Banerjee, Deanna M. Barch, Arielle Baskin-Sommers, Sultana Bhuiyan, Marie-Abele C. Bind, Amanda L. Bogie, Natalie C. Buchbinder, Elliott Bueler, Hülya Bükülmez, B.J. Casey, Linda Chang, Duncan B. Clark, Rebecca G. Clifton, Katharine N. Clouser, Lesley Cottrell, Kelly Cowan, Viren D’Sa, Mirella Dapretto, Soham Dasgupta, Walter Dehority, Kirsten B. Dummer, Matthew D. Elias, Shari Esquenazi-Karonika, Danielle N. Evans, E. Vincent S. Faustino, Alexander G. Fiks, Daniel Forsha, John J. Foxe, Naomi P. Friedman, Greta Fry, Sunanda Gaur, Dylan G. Gee, Kevin M. Gray, Ashraf S. Harahsheh, Andrew C. Heath, Mary M. Heitzeg, Christina M. Hester, Sophia Hill, Laura Hobart-Porter, Travis K.F. Hong, Carol R. Horowitz, Daniel S. Hsia, Matthew Huentelman, Kathy D. Hummel, William G. Iacono, Katherine Irby, Joanna Jacobus, Vanessa L. Jacoby, Pei-Ni Jone, David C. Kaelber, Tyler J. Kasmarcak, Matthew J. Kluko, Jessica S. Kosut, Angela R. Laird, Jeremy Landeo-Gutierrez, Sean M. Lang, Christine L. Larson, Peter Paul C. Lim, Krista M. Lisdahl, Brian W. McCrindle, Russell J. McCulloh, Alan L. Mendelsohn, Torri D. Metz, Lerraughn M. Morgan, Eva M. Müller-Oehring, Erica R. Nahin, Michael C. Neale, Manette Ness-Cochinwala, Sheila M. Nolan, Carlos R. Oliveira, Matthew E. Oster, R. Mark Payne, Hengameh Raissy, Isabelle G. Randall, Suchitra Rao, Harrison T. Reeder, Johana M. Rosas, Mark W. Russell, Arash A. Sabati, Yamuna Sanil, Alice I. Sato, Michael S. Schechter, Rangaraj Selvarangan, Divya Shakti, Kavita Sharma, Lindsay M. Squeglia, Michelle D. Stevenson, Jacqueline Szmuszkovicz, Maria M. Talavera-Barber, Ronald J. Teufel, Deepika Thacker, Mmekom M. Udosen, Megan R. Warner, Sara E. Watson, Alan Werzberger, Jordan C. Weyer, Marion J. Wood, H. Shonna Yin, William T. Zempsky, Emily Zimmerman, Benard P. Dreyer, the RECOVER Initiative

**Affiliations:** Department of Pediatrics, New York University Grossman School of Medicine, New York, NY, USA; Department of Biostatistics, Massachusetts General Hospital, Boston, MA, USA; Department of Pediatrics, Columbia University Vagelos College of Physicians and Surgeons, New York, NY, USA; Department of Laboratory Medicine and Pathology, Mayo Clinic Hospital, Rochester, MN, USA; Avera Research Institute, Avera Health, Sioux Falls, SD, USA; Department of Pediatrics, University of California San Francisco, San Francisco, CA, USA; Department of Sociology, Lamar University, Beaumont, TX, USA; Department of Child and Adolescent Psychiatry, New York University Grossman School of Medicine, New York, NY, USA; Public Health Research Institute and Department of Medicine, Rutgers New Jersey Medical School, Newark, NJ, USA; Center for Human Development, Cognitive Science, Psychiatry, Radiology, University of California San Diego, La Jolla, CA, USA; Department of Medicine, Harvard Medical School, Boston, MA, USA; Department of Medicine, New York University Grossman School of Medicine, New York, NY, USA; Department of Physical Medicine and Rehabilitation, Virginia Commonwealth University School of Nursing, Richmond, VA, USA; Department of Pediatrics, Division of Population Health, Quality, and Implementation Sciences (POPQuIS), Rutgers Robert Wood Johnson Medical School, New Brunswick, NJ, USA; Department of Pediatrics, Columbia University Medical Center: Columbia University Irving Medical Center, New York, NY, USA; Department of Infectious Diseases, Children’s Hospital Los Angeles and the Keck School of Medicine, University of Southern California, Los Angeles, CA, USA; Department of Population Health, New York University Grossman School of Medicine, New York, NY, USA; Department of Cardiology, Boston Children’s Hospital, Boston, MA, USA; Department of Pediatrics, University of California San Diego School of Medicine, San Diego, CA, USA; School of Nursing, Virginia Commonwealth University, Richmond, VA, USA; Departments of Pediatrics and Biostatistics, University of Arkansas for Medical Sciences, Little Rock, AR, USA; Department of Child and Adolescent Psychiatry, Hassenfeld Children’s Hospital at NYU Langone, New York, NY, USA; Department of Pediatrics, Division of Child and Adolescent Health, Columbia University Vagelos College of Physicians and Surgeons and NewYork-Presbyterian, New York, NY, USA; Division of Pediatric Respiratory Medicine, University of California San Diego, San Diego, CA, USA; Division of Pediatric Cardiology, University of Utah and Primary Children’s Hospital, Salt Lake City, UT, USA; Department of Pediatrics, Children’s Hospital Los Angeles, Los Angeles, CA, USA; Department of Pediatrics and Radiology, Children’s Hospital Los Angeles, Los Angeles, CA, USA; Department of Pulmonary Research, Rady Children’s Hospital-San Diego, San Diego, CA, USA; College of Engineering, Northeastern University, Boston, MA, USA; Division of Pediatric Cardiology, NewYork-Presbyterian/Columbia University Irving Medical Center, New York, NY, USA; Department of Pediatrics, Hackensack University Medical Center, Hackensack, NJ, USA; Department of Pediatrics, Medical University of South Carolina, Charleston, SC, USA; Oxley College of Health Sciences, Laureate Institute for Brain Research, Tulsa, OK, USA; Center for Health Sciences, SRI International, Menlo Park, CA, USA; Department of Pediatrics, Kapiolani Medical Center for Women and Children, Honolulu, HI, USA; Department of Pathology and Laboratory Medicine, Children’s Mercy Hospital, Kansas City, MO, USA; Department of Psychological & Brain Sciences, Psychiatry, and Radiology, Washington University in St. Louis, Saint Louis, MO, USA; Department of Psychology, Yale University, New Haven, CT, USA; Department of Pediatrics, University of Oklahoma Health Science Center, Oklahoma City, OK, USA; Center for Human Development, University of California San Diego, San Diego, CA, USA; Department of Pediatrics, Division of Rheumatology, The MetroHealth System, Case Western Reserve University, Cleveland, OH, USA; Department of Neuroscience and Behavior, Barnard College - Columbia University, New York, NY, USA; Department of Diagnostic Radiology and Nuclear Medicine, University of Maryland School of Medicine, Baltimore, MD, USA; Departments of Psychiatry and Pharmaceutical Sciences, University of Pittsburgh, Pittsburgh, PA, USA; Biostatistics Center, George Washington University, Washington, DC, USA; Department of Pediatrics, Hackensack Meridian School of Medicine, Nutley, NJ, USA; Department of Pediatrics, West Virginia University, Morgantown, WV, USA; Department of Pediatrics, Robert Larner M.D. College of Medicine at the University of Vermont, Burlington, VT, USA; Department of Pediatrics, Rhode Island Hospital, Providence, RI, USA; Department of Psychiatry and Biobehavioral Sciences, University of California Los Angeles, Los Angeles, CA, USA; Department of Pediatrics, Norton Children’s Hospital, University of Louisville, Louisville, KY, USA; Department of Pediatrics, Division of Infectious Diseases, University of New Mexico, Albuquerque, NM, USA; Department of Pediatrics, University of California San Diego, San Diego, CA, USA; Division of Cardiology, Children’s Hospital of Philadelphia, Philadelphia, PA, USA; Arkansas Children’s Research Institute, Arkansas Children’s Hospital, Little Rock, AR, USA; Department of Pediatrics, Yale School of Medicine, New Haven, CT, USA; Department of Pediatrics, Children’s Hospital of Philadelphia, Philadelphia, PA, USA; Department of Cardiology, Children’s Mercy Kansas City, Ward Family Heart Center, Kansas City, MO, USA, Kansas City, MO, USA; Department of Neuroscience, University of Rochester School of Medicine and Dentistry, Rochester, NY, USA; Institute for Behavioral Genetics and Department of Psychology and Neuroscience, University of Colorado Boulder, Bolder, CO, USA; Pennington Biomedical Research Center Clinic, Pennington Biomedical Research Center, Baton Rouge, LA, USA; Department of Pediatrics, Rutgers Robert Wood Johnson Medical School, New Brunswick, NJ, USA; Department of Psychiatry and Behavioral Sciences, Medical University of South Carolina, Charleston, SC, USA; Department of Pediatrics, Division of Cardiology, George Washington University School of Medicine & Health Sciences, Washington, DC, USA; Department of Psychiatry, Washington University School of Medicine, St Louis, MO, USA; Department of Psychiatry, University of Michigan, Ann Arbor, MI, USA; Division of Practice-Based Research, Innovation, & Evaluation, American Academy of Family Physicians, Leawood, KS, USA; Departments of Pediatrics and Physical Medicine & Rehabilitation, Section of Pediatric Rehabilitation, University of Arkansas for Medical Sciences, Little Rock, AR, USA; Center for Health Equity and Community Engaged Research and Department of Population Health Science and Policy, New York, NY, USA; Clinical Trials Unit, Pennington Biomedical Research Center, Baton Rouge, LA, USA; Division of Neurogenomics, Translational Genomics Research Institute, Phoenix, AZ, USA; Department of Pediatrics, University of Arkansas for Medical Sciences, Little Rock, AR, USA; Department of Psychology, University of Minnesota, Minneapolis, MN, USA; Department of Pediatrics, Arkansas Children’s Hospital, University of Arkansas Medical School, Little Rock, AR, USA; Department of Psychiatry, University of California San Diego, La Jolla, CA, USA; Department of Obstetrics, Gynecology, and Reproductive Sciences, University of California San Francisco, San Francisco, CA, USA; Department of Pediatrics, Pediatric Cardiology, Lurie Children’s Hospital, Northwestern University Feinberg School of Medicine, Chicago, IL, USA; Departments of Pediatrics, Internal Medicine, and Population and Quantitative Health Sciences, Case Western Reserve University, Cleveland, OH, USA; Department of Pediatric Clinical Research, Medical University of South Carolina, Charleston, SC, USA; Department of Physics, Florida International University, Miami, FL, USA; Department of Pediatrics, Respiratory Medicine Division, University of California San Diego, San Diego, CA, USA; Heart Institute, Cincinnati Children’s Hospital Medical Center, Cincinnati, OH, USA; Department of Psychology, University of Wisconsin-Milwaukee, Milwaukee, WI, USA; Department of Pediatric Infectious Disease, Avera McKennan University Health Center, University of South Dakota, Sioux Falls, SD, USA; Department of Pediatrics, University of Toronto, Labatt Family Heart Center, The Hospital for Sick Children, Toronto, ON, Canada; Department of Pediatrics, University of Nebraska Medical Center, Omaha, NE, USA; Department of Pediatrics, Division of Developmental-Behavioral Pediatrics, New York University Grossman School of Medicine, New York, NY, USA; Department of Obstetrics and Gynecology, University of Utah Health, Salt Lake City, UT, USA; Department of Pediatrics, Valley Children’s Healthcare, Department of Pediatrics, Madera, CA, Madera, CA, USA; Department of Psychiatry, Virginia Commonwealth University, Richmond, VA, USA; Department of Pediatrics, New York Medical College, Valhalla, NY, USA; Department of Pediatrics, Section of Infectious Diseases and Global Health, Yale University School of Medicine, New Haven, CT, USA; Department of Pediatric Cardiology, Children’s Healthcare of Atlanta, Atlanta, GA, USA; Department of Pediatrics, Division of Pediatric Cardiology, Riley Hospital for Children, Indiana University School of Medicine, Indianapolis, IN, USA; Department of Pediatrics, University of New Mexico, Health Sciences Center, Albuquerque, NM, USA; Department of Pediatrics, Division of Infectious Diseases, Epidemiology and Hospital Medicine, University of Colorado Anschutz Medical Campus, Aurora, CO, USA; Department of Pediatrics, University of Michigan Health System, Ann Arbor, MI, USA; Department of Pediatric Cardiology, Phoenix Children’s Hospital, Phoenix, AZ, USA; Division of Pediatric Cardiology, Children’s Hospital of Michigan, Detroit, MI, USA; Department of Pediatric Infectious Disease, University of Nebraska Medical Center, Omaha, NE, USA; Department of Pediatrics, Children’s Hospital of Richmond at Virginia Commonwealth University, Richmond, VA, USA; Department of Pediatrics, Pediatric Cardiology, University of Mississippi Medical Center, Jackson, MS, USA; Department of Pediatrics, University of Texas Southwestern Medical Center, Dallas, TX, USA; Department of Pediatrics, University of Louisville School of Medicine, Louisville, KY, USA; Division of Cardiology, Children’s Hospital Los Angeles, Los Angeles, CA, USA; Department of Pediatrics, Avera McKennan Hospital and University Health Center, Sioux Falls, SD, USA; Nemours Cardiac Center, Nemours Childrens Health, Delaware, Wilmington, DE, USA; RECOVER Neurocognitive and Wellbeing/Mental Health Team, NYU Grossman School of Medicine, New York, NY, USA; Center for Individualized Medicine, Mayo Clinic Hospital, Rochester, MN, USA; Departments of Pediatrics and Population Health, New York University Grossman School of Medicine, New York, NY, USA; Department of Pediatrics, Connecticut Children’s Medical Center, Hartford, CT, USA; Department of Communication Sciences & Disorders, Northeastern University, Boston, MA, USA

**Author notes:** These authors contributed equally to this work (Rachel Gross, Tanayott Thaweethai). Membership of the RECOVER Initiative is provided in the Acknowledgements.

## Abstract

**Importance:** The prevalence, pathophysiology, and long-term outcomes of COVID-19 (post-acute sequelae of SARS-CoV-2 [PASC] or “Long COVID”) in children and young adults remain unknown. Studies must address the urgent need to define PASC, its mechanisms, and potential treatment targets in children and young adults.

**Observations:** We describe the protocol for the Pediatric Observational Cohort Study of the NIH’s **RE**searching **COV**ID to **E**nhance **R**ecovery (RECOVER) Initiative. RECOVER-Pediatrics is an observational meta-cohort study of caregiver-child pairs (birth through 17 years) and young adults (18 through 25 years), recruited from more than 100 sites across the US. This report focuses on two of five cohorts that comprise RECOVER-Pediatrics: 1) a *de novo* RECOVER prospective cohort of children and young adults with and without previous or current infection; and 2) an extant cohort derived from the Adolescent Brain Cognitive Development (ABCD) study (*n*=10,000). The *de novo* cohort incorporates three tiers of data collection: 1) remote baseline assessments (Tier 1, n=6000); 2) longitudinal follow-up for up to 4 years (Tier 2, n=6000); and 3) a subset of participants, primarily the most severely affected by PASC, who will undergo deep phenotyping to explore PASC pathophysiology (Tier 3, n=600). Youth enrolled in the ABCD study participate in Tier 1. The pediatric protocol was developed as a collaborative partnership of investigators, patients, researchers, clinicians, community partners, and federal partners, intentionally promoting inclusivity and diversity. The protocol is adaptive to facilitate responses to emerging science.

**Conclusions and Relevance:** RECOVER-Pediatrics seeks to characterize the clinical course, underlying mechanisms, and long-term effects of PASC from birth through 25 years old. RECOVER-Pediatrics is designed to elucidate the epidemiology, four-year clinical course, and sociodemographic correlates of pediatric PASC. The data and biosamples will allow examination of mechanistic hypotheses and biomarkers, thus providing insights into potential therapeutic interventions.

**Clinical Trials.gov Identifier:** Clinical Trial Registration: http://www.clinicaltrials.gov. Unique identifier: NCT05172011

## Introduction

The coronavirus disease 2019 (COVID-19) pandemic has significantly impacted child health. Nearly 100 million people have been diagnosed with COVID-19 in the United States (US), with nearly 16 million children [1]. Although it is estimated that between 10% and 30% of adults experience persistent symptoms from COVID-19 [2], termed post-acute sequelae of SARS-CoV-2 (PASC) or Long COVID, the prevalence in children is less well-established [3, 4]. As an emerging illness, the absence of universally-accepted PASC definitions in children challenge the elucidation of its epidemiology.

Unique challenges in understanding PASC symptoms in children have likely contributed to the limited evidence. Given that young children might not be able to articulate symptoms, studies must rely on caregiver interpretation. In addition, manifestation of symptoms may vary substantively across stages of physiological, emotional, and cognitive development [5]. As the medical community shifts from managing serious acute disease to addressing long-term consequences, large scale studies are needed to define and recognize PASC in children across the life course, to understand its natural history, and to develop evidence to guide successful treatment.

The pandemic began with a misconception that children were spared [6]. We now recognize that children and families are greatly impacted during both acute and chronic phases [7–10]. One distinct manifestation in children was recognized in April 2020; now called Multisystem Inflammatory Syndrome in Children (MIS-C) [11]. This debilitating hyperinflammatory syndrome has impacted over 9,000 children and young adults in the US [12], and represents a distinct post-acute syndrome that is typically recognizable in clinical practice. Other more chronic manifestations of PASC are challenging to characterize and identify. Furthermore, children with PASC may present with different symptoms and greater mental health concerns than adults [1, 13–19]. Additional phenotypes of childhood PASC are being reported, including phenotypes similar to postural orthostatic tachycardia syndrome (POTS), myalgic encephalomyelitis/chronic fatigue syndrome (ME/CFS), postintensive care unit syndrome, and potentially many others [20–22]. Therefore, a compelling rationale exists to invest resources and effort to study PASC in children. The NIH’s REsearching COVID to Enhance Recovery (RECOVER) Initiative responded by bringing together researchers, communities, and families in a systematic study of PASC in children [23]. Evidence that leads to improved health trajectories of children with PASC, could have population-level health impacts for decades to come.

### Study rationale

RECOVER has established the Pediatric Observational Cohort Study (RECOVER-Pediatrics), which is a combined retrospective and prospective longitudinal study, including five distinct cohorts, integrated together as a meta-cohort [23]. The overall goal is to characterize the clinical course, underlying mechanisms and long-term health effects of PASC on children and young adults from birth through 25 years old, to inform future pediatric preventive and treatment measures.

### Study aims

RECOVER-Pediatrics scientific aims are to:

1. Characterize the prevalence and incidence of new onset or worsening symptoms related to PASC
2. Characterize the spectrum of clinical symptoms of PASC, including distinct phenotypes, and describe the clinical course and recovery.
3. Identify risk and resiliency factors for developing PASC and recovering from PASC.
4. Define the pathophysiology of PASC, including subclinical organ dysfunction, and identify biological mechanisms underlying the pathogenesis of PASC.

## Materials and Methods

### Overview of study design

RECOVER-Pediatrics is a longitudinal, observational meta-cohort study of children and young adults (ages birth through 25 years) and their caregivers, recruited from healthcare- and community-based settings in more than 100 sites throughout the US, including Puerto Rico. Those *with and without* a history of a SAR-CoV-2 infection are included. For those 17 years or younger, data are collected by caregiver report and child direct assessments, and for those 18 through 25 years old by self-report. The study is being conducted from March 2022 to March 2026.

The pediatric meta-cohort is comprised of five distinct cohorts: 1) *de novo RECOVER prospective cohort* including children and young adults ages birth through 25 years, with or without a known history of infection, and their caregivers; 2) *Adolescent Brain Cognitive Development (ABCD)* extant cohort, the largest long-term US study of brain development in adolescence [24, 25]; 3) *In utero exposure cohort*, including children less than 3 years old born to individuals with and without a SAR-CoV-2 infection during pregnancy [26, 27]; 4) *COVID MUSIC Study* extant cohort (*Long-Term Outcomes after the Multisystem Inflammatory Syndrome In Children*), including children and young adults with history of MIS-C [28]; and 5) cohort of children and young adults with a history of *post-COVID vaccine myocarditis*. This report focuses on the *de novo* cohort and ABCD (**Figure 1**).

**Fig 1:**
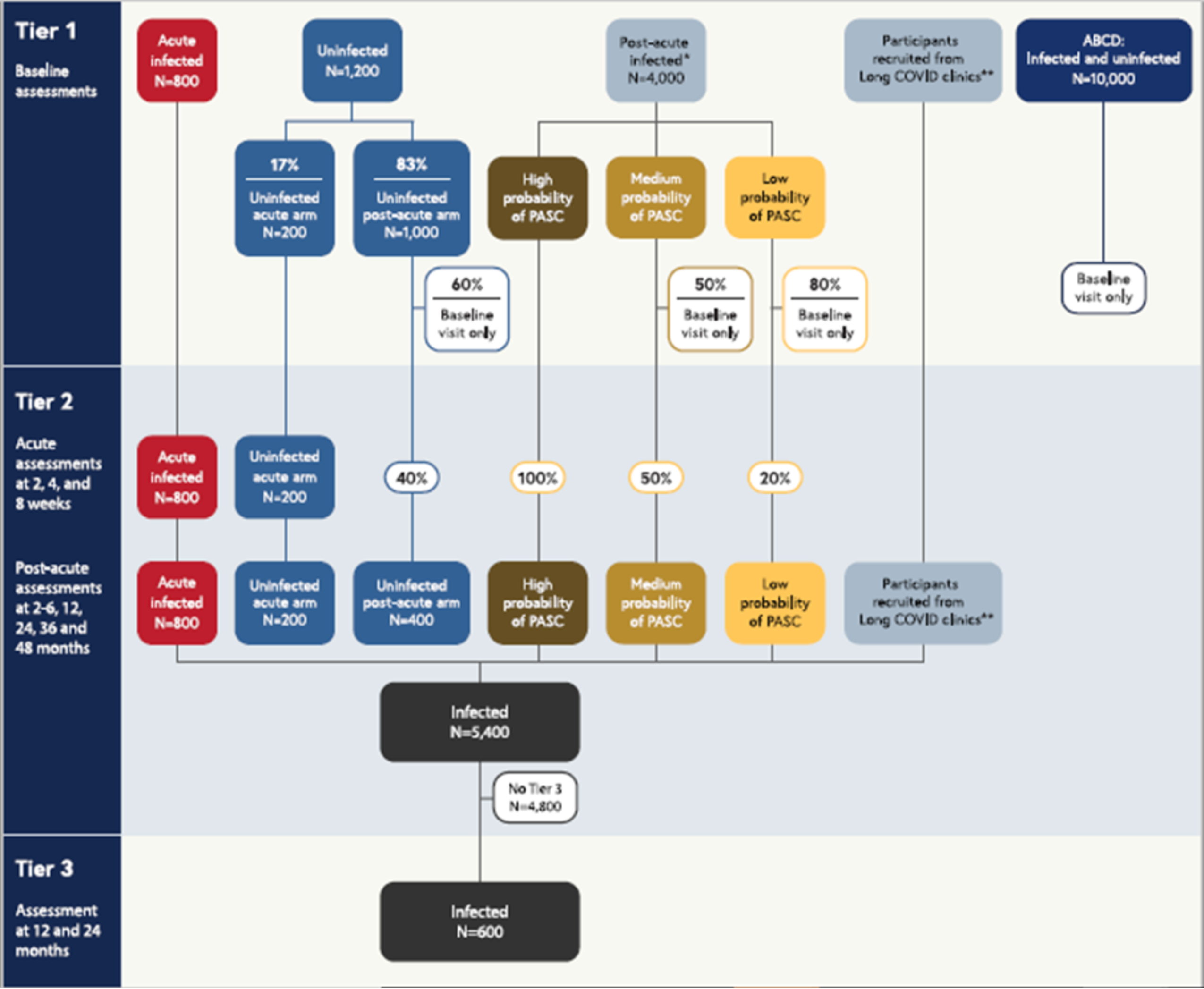
Overview of RECOVER-Pediatrics (de novo and ABCD cohorts).

Figure 1 shows a tiered overview of 2 of the 5 cohorts included in the meta-cohort (*de novo* RECOVER prospective cohort and ABCD), their participation in the three study tiers, and their targeted sample sizes (see *Study Participants*). Children and young adults ages newborn through 25 years old will be enrolled in the meta-cohort at Tier 1 for the *de novo* RECOVER prospective cohort (more than 6,000 from birth through 25 years old, including those with and without history of infection), and from ABCD (up to 10,000 adolescents with and without history of infection). All children and young adults enrolled in the study complete a baseline assessment (Tier 1). Percentages shown indicate random sampling proportions. Children and young adults without history of infection are assigned at random with prespecified proportions to the acute and post-acute arms. All children and young adults with history of infection who enroll into the acute arm and those without a history of infection who are randomized to the acute arm are asked to complete assessments at 2, 4, and 8 weeks. Following a promotion algorithm (Table 2), children and young adults in Tier 1 will be selected to be promoted to Tier 2, which includes assessments at 2-6, 12, 24, 36, and 48 months after enrollment. 600 children and young adults with history of infection, selected from Tier 2, will complete more intensive Tier 3 assessments at 12 and 24 months after enrollment.

*Children and young adults with history of infection who enroll in the post-acute arm (“post-acute infected”, n=4,000) are stratified into High, Medium, and Low probability of PASC groups based on a combination of past Long COVID diagnoses, Tier 1 Global PROMIS health measures, and symptom survey screener responses. Then, 100% of the high probability group, 50% of the medium probability group, and 20% of the low probability group are promoted at random to Tier 2. Since the distribution of these probability groups is unknown a priori, sample sizes are not specified for each category. Overall, the number of children and young adults who progress to Tier 2 will be less than the initial post-acute infected sample size, but the total target sample size for infected children and young adults in Tier 2 is 5,400.

** In order to achieve a sample of 5,400 children and young adults with history of infection in Tier 2 that is skewed towards those with greater likelihood of having PASC, additional children and young adults will be recruited from Long COVID clinics and subspecialty services to complete both Tier 1 and Tier 2 assessments.

RECOVER-Pediatrics is structured in a sequential fashion with three Tiers of data collection. Participants may be enrolled initially into Tier 1, which consists of a broad screening of health using remote surveys and biospecimen collection. Participants may subsequently progress to Tier 2, which includes a detailed review of health collected longitudinally for up to four years, using a combination of remote surveys and in-person assessments of biological and psychosocial data. In order to achieve the sample size required for Tier 2 assessments, other study participants will be recruited who present with a high probability of having PASC, such as those directly recruited from a clinic that focuses on Long COVID or presenting with a physician diagnosis of PASC. These participants will receive Tier 1 assessments and progress directly to Tier 2. Finally, in Tier 3, a subset of children and young adults most severely affected by PASC will undergo deep phenotyping with more intensive assessments to study PASC pathophysiology.

RECOVER-Pediatrics Tier 1 assessments aim to characterize the prevalence and incidence of new onset or worsening of sustained COVID-related symptoms (aim 1) and to gain a comprehensive understanding of the impact of exposure to a SARS-CoV-2 infection on broad physical, behavioral and mental health (aim 2). Tier 2 facilitates studying the natural history of PASC symptoms and potential recovery over time (aim 2). Child, household, and caregiver factors gathered in Tier 1, such as social determinants of health and prior health conditions, will be assessed to determine how they increase the risk of or protect against specific clinical outcomes (aim 3). Finally, Tier 3 data investigates long-term effects on multiple organ systems and child development (aim 4). Additionally, integration of Tier 1 and Tier 2 data will allow investigation of COVID-disease exposures and experiences which may be responsible for the clinical patterns observed in Tier 3.

The pediatric protocol was designed through collaboration across key stakeholders, including patients, caregivers, researchers, clinicians, community partners, and federal partners, fostering a patient-centered approach and promoting inclusivity and diversity. The pediatric protocol is adaptive to facilitate the changes needed in light of emerging science and the evolving pandemic.

### Study organizational structure and management

Study infrastructure includes four cores: 1) Clinical Science Core (CSC) at the NYU Grossman School of Medicine, which oversees study sites and provides scientific leadership in collaboration with hub and site Principal Investigators; 2) Data Resource Core (DRC) at Massachusetts General Hospital and Brigham and Women’s Hospital, which provides scientific and statistical leadership, and handles data management and storage; 3) PASC Biorepository Core (PBC) at Mayo Clinic, which manages biospecimens obtained; and 4) Administrative Coordinating Center (ACC) at RTI International, which provides operational and administrative support; collectively these form the Core Operations Group. The four cores are supported by oversight committees and pathobiology task forces provide content-specific input. RECOVER cohort studies are overseen by the National Community Engagement Group (NCEG) composed of patient and community representatives, a Steering Committee composed of site Principal Investigators and NIH program leadership, an Executive Committee composed of NIH Institute leadership, and an Observational Safety Monitoring Board composed of experts in longitudinal observational studies, epidemiology, bioethics, and biostatistics. RECOVER-Pediatrics includes 10 hubs that manage ∼100 sites (**Supplemental Table 1**). NYU Grossman School of Medicine serves as the sIRB for sites without prior reliance agreements.

### Recruitment, consent, and screening strategies

The *de novo RECOVER prospective cohort study* is recruiting participants from healthcare- and community-based settings. Healthcare-based recruitment involves local media, text messaging, hospital websites, COVID registries, and partnerships with pediatric practices, nurse hotlines, or emergency departments. Community-based recruitment includes partnering with community health workers, school nurses, sports coaches, health fairs, and a mobile van to access rural communities. Participants can also join by self-referral through the RECOVER website, or in response to plain language and picture-based recruitment materials in both English and Spanish, which were developed with community input and using health literacy principles [29].

Eligible dyads complete an informed consent or assent process at enrollment for Tiers 1 and 2 (in-person or via electronic informed consent [e-consenting]). Young adults, aged 18 through 25 years old, sign their own informed consent. Tier 3 consent forms will only be completed when testing is offered.

In ABCD, 11,880 children aged 9-10 years old were recruited from community and school sites to participate in a 10-year study with the goal of understanding neurocognitive development during adolescence [24, 25, 30]. All ABCD participants are being contacted and offered enrollment into RECOVER-Pediatrics.

### Eligibility criteria

Children and young adults from birth through 25 years old are eligible to be enrolled in the *de novo cohort*, regardless of history of SARS CoV-2 infection. Enrolled participants are then categorized as either “infected” or “uninfected”: Infected participants have history of suspected, probable, or confirmed SARS-CoV-2 infection, defined by the World Health Organization (WHO) criteria [31], evidence of infection by serum antibody profile, or a history of MIS-C. Uninfected participants are those who self-report as having no history of a SARS-CoV-2 infection and who have never met WHO criteria; they have no evidence of a past asymptomatic infection in their medical history or evidence of past infection by serum antibody profile.

A primary caregiver, defined as an individual responsible for the enrolled child or young adult who resides in the same household, such as biological or nonbiological family member, is invited to enroll.

The primary exclusion criterion is any child or young adult with co-morbid illness with expected survival of less than 2 years. There is no limit to the number of children or young adults who can be enrolled from a single household. See supplemental tables for detailed eligibility criteria, definitions of analytic groups, and the World Health Organization Criteria (**Supplement Tables 2, 3, and 4**, respectively).

### Study participants

Recruitment is striving for a diverse sample that generally represents the US population, and encourages participation from rural or medically underserved communities, non-English speaking participants, and non-hospitalized participants with an acute COVID-19 infection. Participants are compensated for completing assessments and reimbursed for excess travel.

At least 6,000 participants will be recruited into the *de novo* cohort (**Figure 1**). Children and young adults with history of infection are classified into *one of two study arms* (acute arm vs. post-acute arm), based on their history of SARS-CoV-2 infection and infection dates. The *acute arm* includes 800 children and young adults whose most recent SARS-CoV-2 infection was 30 days or less prior to enrollment. The *post-acute arm* includes 4,000 children and young adults whose most recent SARS-CoV-2 infection was greater than 30 days prior to enrollment. In the group without a history of infection, 1,200 children and young adults will be randomly assigned to follow either the acute (200, or 17%) or post-acute (1,000 or 83%) arm of the protocol. Additional children and young adults will be recruited from Long COVID clinics and other subspecialty services in order to achieve Tier 2 sample size targets (see *Timing of Study Assessments*).

Up to 10,000 participants will also be recruited from the ABCD cohort.

### Timing of study assessments

The assessments for the *de novo cohort* consists of three tiers, which vary in timing, collection methods and intensity.

*Tier 1* (baseline visit for all participants) includes a single visit that is completed either via self-administration (remote and electronic) or research staff-assisted collection (e.g., telephone, videoconference, or in-person).

*Tier 2* (follow-up visits) includes five longitudinal in-person visits at 2 to 6-, 12-, 24-, 36- and 48-months post-enrollment. The children and young adults followed longitudinally in Tier 2 are selected based on a sampling scheme that prioritizes the acute arm as well as children and youth in the post-acute arm with a greater likelihood of having PASC. Promotion to Tier 2 occurs as follows: 1) All children/young adults in the *acute arm* with or without history of infection will be promoted; 2) children/young adults in the *post-acute arm with a history of infection* will be promoted at a rate dependent on their likelihood of PASC based on prior Long COVID diagnoses, Tier 1 PROMIS global health measure responses [32–34], and symptoms screener survey responses [16, 35] (**Table 1**); and 3) 40% of children/young adults without known infection in the post-acute arm, selected at random, will be promoted. In addition to promoting children and young adults from Tier 1, children and young adults will also be recruited from Long COVID clinics and subspecialty services to achieve the target sample size in Tier 2 of 6,000. These children and young adults will complete both Tier 1 and Tier 2 assessments. See **Table 2** for a full description of the promotion algorithm.

**Table 1:**
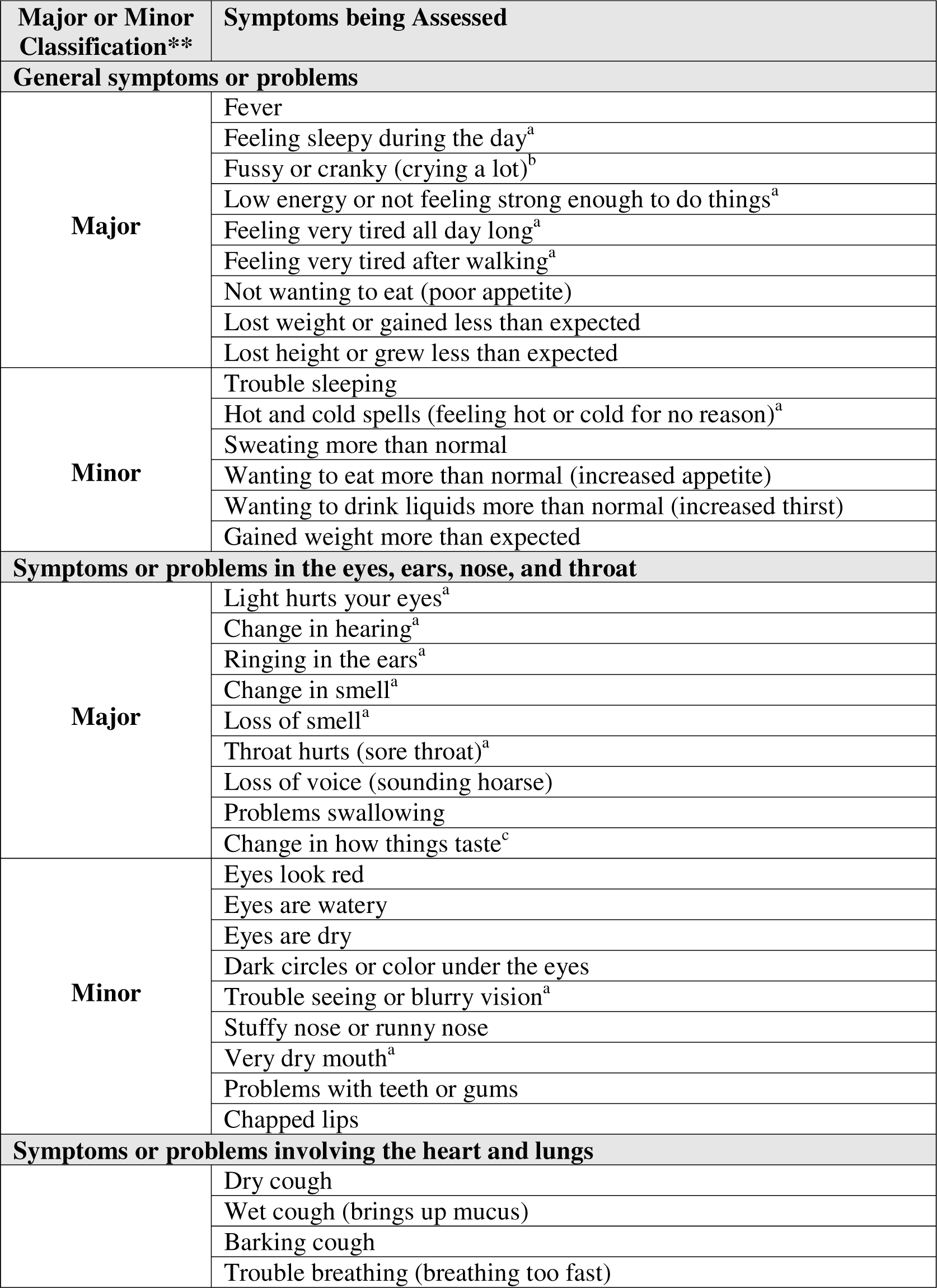
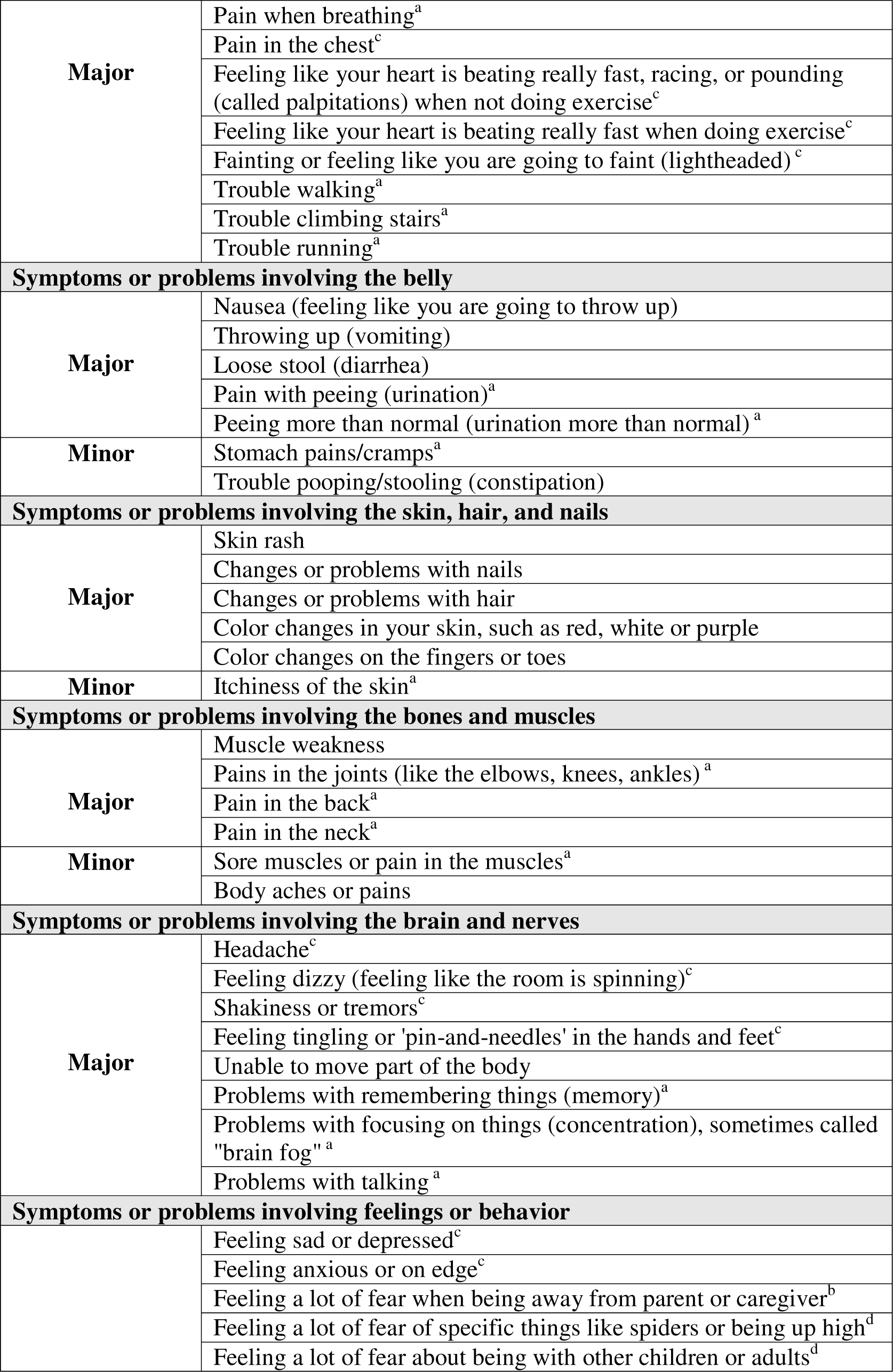
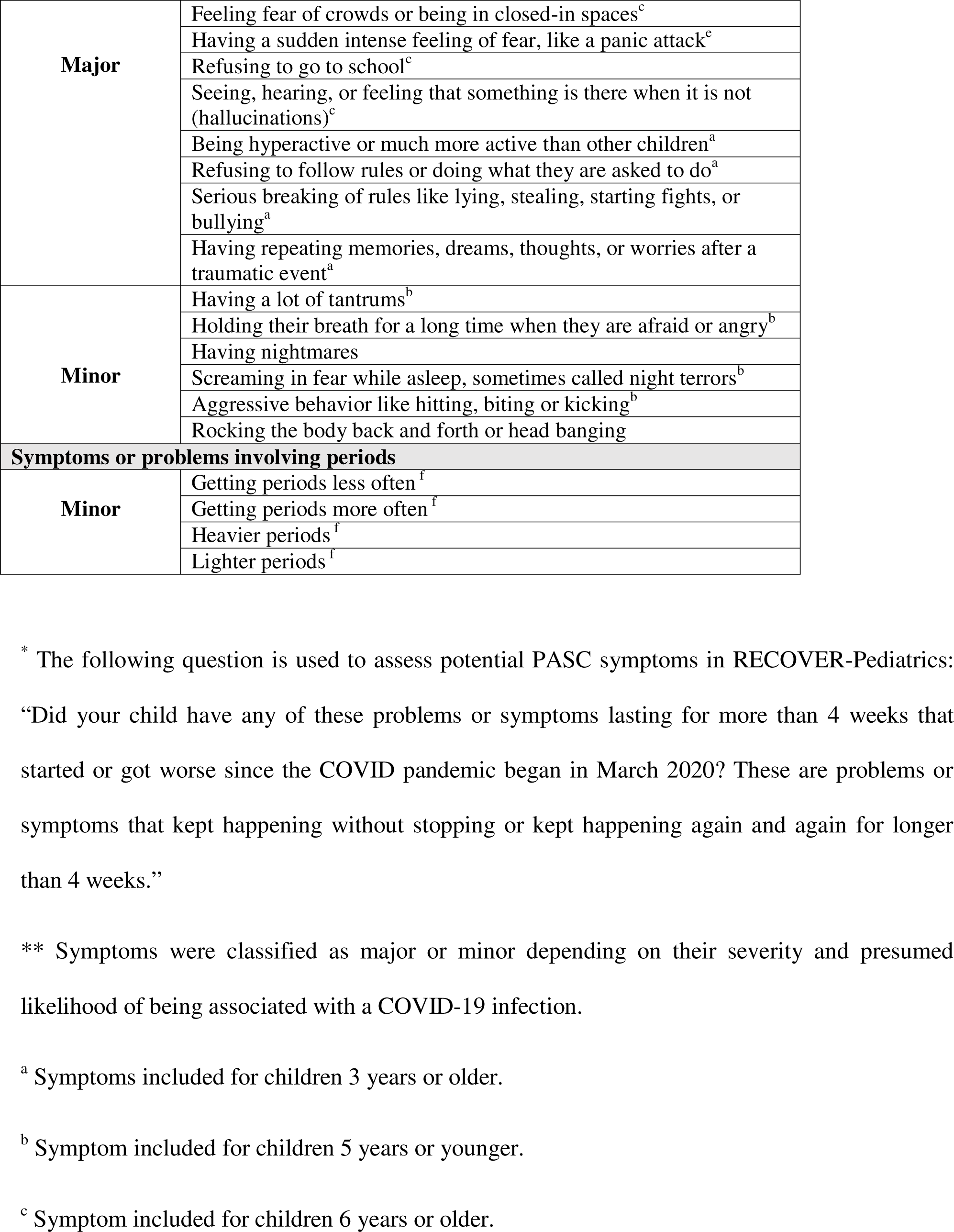
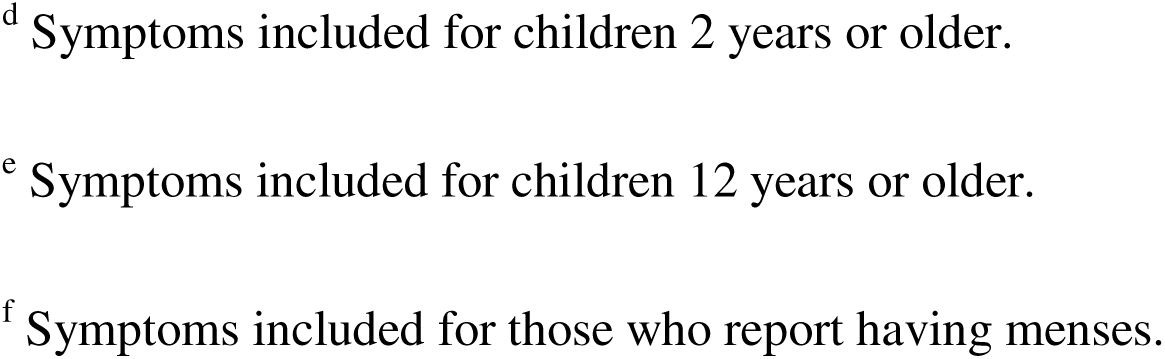
Potential Post-Acute Sequelae of SARS-CoV-2 (PASC) Symptoms Being Assessed in RECOVER-Pediatrics*.

**Table 2.** Promotion Algorithm Used in the *de novo* RECOVER-Pediatrics Cohort for Selecting Children and Young Adults for the Longitudinal Follow-Up (Tier 2).

| Type of participant | Subgroup | Subgroup criteria | Promotion rate to Tier 2 |
| --- | --- | --- | --- |
| “Acute infected”: Children and young adults who reported having a COVID infection $\leq 30$ days prior to enrollment | All | None | 100% |
| “Post-acute infected”: Children and young adults who reported having a COVID infection $> 30$ days prior to enrollment | High probability of PASC<sup>a</sup> | Any of the following:<br>1. Prior diagnosis of Long COVID or MIS-C based on study site medical record review or referral by a health care provider<br>2. Recruitment from a Long COVID clinic<br>3. $\geq 1$ fair/poor responses on the global health PROMIS scale<sup>b</sup> and $\geq 1$ major or minor symptom reported<sup>c</sup><br>4. $\geq 1$ good/fair/poor responses on the global health PROMIS scale<sup>b</sup> and $\geq 1$ major symptom reported<sup>c</sup><br>5. $\geq 1$ good/fair/poor responses on the global health PROMIS scale<sup>b</sup> and $\geq 2$ minor symptoms reported<sup>c</sup> | 100% |
|  | Medium probability of PASC<sup>a</sup> | Any of the following:<br>1. $\geq 1$ good responses on the global health PROMIS scale<sup>b</sup> and $\geq 1$ major or minor symptom reported<sup>c</sup><br>2. $\geq 1$ very good/good/fair/poor responses on the global health PROMIS scale<sup>b</sup> and $\geq 1$ major symptom reported<sup>c</sup><br>3. $\geq 1$ very good/good/fair/poor responses on the global health PROMIS scale<sup>b</sup> and $\geq 2$ minor symptoms reported<sup>c</sup> | 50% |
|  | Low probability of PASC<sup>a</sup> | Does not meet high or medium probability of PASC criteria | 20% |
| “Uninfected”: Children or Young adults | Acute arm | At enrollment, 17% of the “uninfected” group were randomly assigned to participate in the acute arm of the study. | 100% |
| without a known history of a COVID infection | Post-acute arm | At enrollment, 83% of the “uninfected” group were randomly assigned to participate in the post-acute arm of the study. 40% of this group will be randomly assigned to participate in Tier 2 | 40% |

<sup>a</sup> Responses to the PROMIS Global Health Scales and the presence of major and minor symptoms are used to categorize participants who are post-acute infected as high, medium, or low probability of PASC.
<sup>b</sup> The PROMIS Global Health Scales are self-reported or caregiver-reported measures of overall, physical, and mental health for young adults and children, respectively [32-34]. The three questions from the caregiver-reported version that are used in the algorithm, include: 1) “*In general, would you say your child’s health is?*”; 2) “*In general, how would you rate your child’s physical health?*”; and 3) “*In general, how would you rate your child’s mental health, including mood and ability to think?*” Responses include: *Excellent, Very Good, Good, Fair, or Poor*.
<sup>c</sup> A list of all major and minor symptoms, reported at the enrollment visit as part of the symptom screener, is provided in the Table 1 [16, 35]. Not all symptoms are asked of all participants, as many are age-specific (e.g., fewer symptoms assessed for younger children) and sex-specific (e.g., menses related symptoms).

Children and young adults in the acute arm with a history of infection will also complete remote assessments at 2, 4, and 8 weeks after their infection onset, with additional in-person assessments at 8 weeks. Children and young adults in the acute arm without history of infection will complete the same assessments, timed relative to their enrollment date. All ABCD youth are eligible to participate in RECOVER Tier 1, and can be referred to a *de novo* cohort site to participate in Tiers 2 and 3, if geographically feasible.

*Tier 3* has the most clinically intensive assessments with longitudinal in-person visits for a subset at 12 and 24 months post-enrollment. Tier 3 will include 600 children and young adults with history of infection from Tier 2.

### Main categories of data

Data collected for the *de novo* and ABCD cohorts are described below (**Table 3**).

**Table 3:**
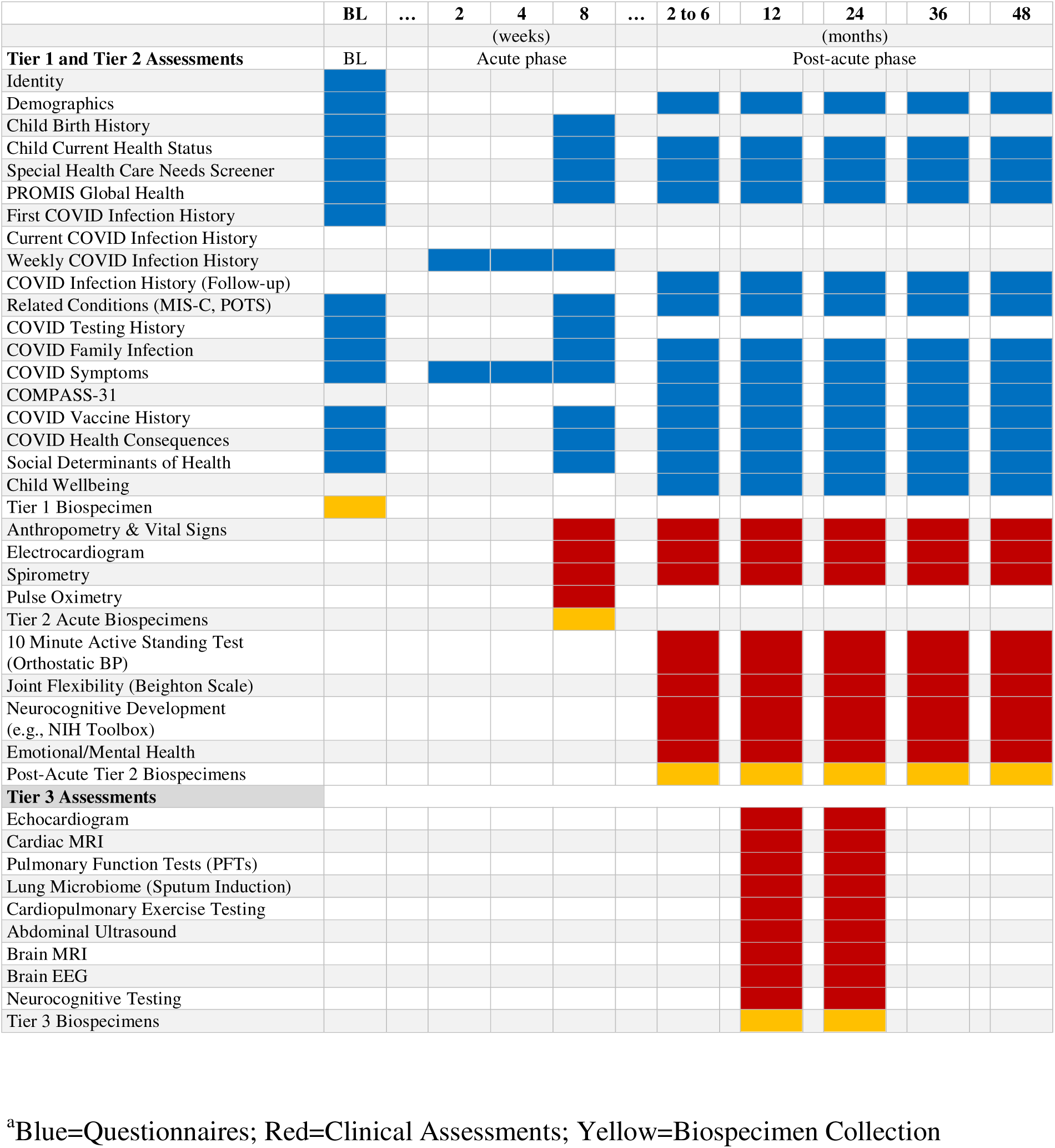
Summary of Study Assessments in the *de novo* RECOVER-Pediatrics Cohort^a^.

*Surveys* include validated surveys with NIH common data elements, as available, informed by expert opinion (**Supplemental Table 5**). All are completed using Research Electronic Data Capture (REDCap), with the child’s first name coded within surveys to personalize the experience and to clarify which child the questions refer to given caregivers can have multiple children enrolled. For youth 17 years or younger, the caregiver is the primary respondent. Participants 18 through 25 years old are the primary respondent. Surveys assess sociodemographic information [36], child birth history [37], special health care needs [37–39], SARS-CoV-2 infection history, related conditions (e.g., MIS-C, POTS or other form of dysautonomia, and Long COVID diagnoses), COVID testing and vaccine history, COVID-related symptoms (both acute and long-term), COVID health consequences (e.g., diet [40], physical activity [40], sleep [40], screen time [40], schooling, parenting [41]) and social determinants of health (e.g., food insecurity [42], social support [43]). A list of potential Long COVID symptoms are assessed [16, 35] (Table 1), with respondents asked whether a specific problem or symptom is/was present for at least 4 weeks since the beginning of the COVID-19 pandemic and, for respondents with a history of infection, if the symptoms started before or after their infection.

*Clinical assessments* are completed at in-person Tier 2 visits across overarching domains of physical growth, physical health, neurocognition, and neurobehavioral function (**Supplemental Table 6**). Physical health domains include anthropometrics, vital signs, an active standing test measuring orthostatic blood pressures [44, 45], joint flexibility tests [46], electrocardiograms, and spirometry. Neurocognitive and neurobehavioral assessments vary by age (**Table 4**).

**Table 4:** Neurocognitive, Neurobehavioral, Well-Being and Mental Health Measures by Age in Tiers 2 and 3 for the *de novo* RECOVER-Pediatrics Cohort.

| **Study Tier** | **Neurocognitive and Developmental Assessments** | **Neurobehavioral, Well-Being and Mental Health Assessments** |
| --- | --- | --- |
| **Infancy and Toddlerhood: Birth through 2 years old** |  |  |
| Tier 2 | Ages and Stages Questionnaire—3rd Edition (ASQ-3) (47-49)<br>Modified Checklist for Autism in Toddlers Revised with Follow up (MCHAT-RF) (50) | Ages and Stages Questionnaires: Social-Emotional, 2nd Edition (ASQ:SE-2) (52)<br>Child Behavior Checklist (53) |
| Tier 3 | N/A | N/A |
| **Preschool-Age: 3 years old through 5 years old** |  |  |
| Tier 2 | Ages and Stages Questionnaire—3rd Edition (ASQ-3) (47-49)<br>NIH Toolbox Cognitive Measures (51) | Ages and Stages Questionnaires: Social-Emotional, 2nd Edition (ASQ:SE-2) (52)<br>Child Behavior Checklist (53)<br>Patient-Reported Outcomes Measurement Information System (PROMIS®) Parent Proxy Anger Scale (54)<br>PROMIS® Parent Proxy Psychological Stress Experiences Scale (55)<br>PROMIS® Parent Proxy Positive Affect Scale (56) |
| Tier 3 | *Cognitive*: Woodcock Johnson Cognitive Battery subtests (60)<br>*Language*: Woodcock-Johnson Oral Language Battery subtests (60)<br>*Verbal Memory*: Woodcock Johnson subtests (60)<br>*Visual Memory*: Wide Range Assessment of Memory and Learning (61)<br>*Visual-Motor Drawing*: Beery Buktenica (62)<br>*Visual Motor Speed*: Purdue Pegboard (63)<br>*Pre-Academics*: Woodcock-Johnson Achievement Battery subtests (60) | Kiddie SADS computer completed by caregiver (64) |
| **School-age and Adolescence: 6 years old through 17 years old** |  |  |
| Tier 2 | NIH Toolbox Cognitive Measures (51) | Patient-Reported Outcomes Measurement Information System (PROMIS®) Parent Proxy Anger Scale (54)<br>PROMIS® Parent Proxy Psychological Stress Experiences Scale (55)<br>PROMIS® Parent Proxy Positive Affect Scale (56)<br>Revised Children’s Anxiety and |
|  |  | Depression Scale (RCADS-25) (57)<br>Strengths and Difficulties Questionnaire (Hyperactivity/Inattention and Conduct Problems Subscales) (58) |
| Tier 3 | *Cognitive:* Woodcock Johnson Cognitive Battery subtests (60)<br>*Language:* Woodcock-Johnson Oral Language Battery subtests (60)<br>*Verbal Memory:* Woodcock Johnson subtests (60)<br>*Visual Memory:* Wide Range Assessment of Memory and Learning (61)<br>*Visual-Motor Drawing:* Beery Buktenica (62)<br>*Visual Motor Speed:* Purdue Pegboard (63)<br>*Pre-Academics:* Woodcock-Johnson Achievement Battery subtests (60) | Kiddie SADS computer completed by caregiver (64) |
| **Young Adults: 18 years through 25 years old** |  |  |
| Tier 2 | NIH Toolbox Cognitive Measures (51) | Patient-Reported Outcomes Measurement Information System (PROMIS®) Parent Proxy Anger Scale (54)<br>PROMIS® Parent Proxy Psychological Stress Experiences Scale (55)<br>Achenbach Adult Self Report (59) |
| Tier 3 | *Cognitive:* Woodcock Johnson Cognitive Battery subtests (60)<br>*Language:* Woodcock-Johnson Oral Language Battery subtests (60)<br>*Verbal Memory:* Woodcock Johnson subtests (60)<br>*Visual Memory:* Wide Range Assessment of Memory and Learning (61)<br>*Visual-Motor Drawing:* Beery Buktenica (62)<br>*Visual Motor Speed:* Purdue Pegboard (63) | SADS Structured Psychiatric Interview (64) |

Neurocognitive domains include broad and specific measures of attention, memory, receptive and expressive language skills, reading, and sensory function [47–51]. Neurobehavioral domains include a broad assessment of behavioral function including anxiety, mood, social interactions, aggression, sleep, self-regulatory behaviors, somatic complaints and attention concerns [52–59]. Tier 3 assessments follow the same domains, but provide more in-depth measurements. The promotion algorithm for Tier 3 is still under development. Physical health domains of cardio-pulmonary function are assessed by echocardiogram, cardiopulmonary exercise testing, cardiac MRI, pulmonary function tests, and sputum induction. Gastrointestinal function is assessed using abdominal ultrasound, and neurological function is assessed using brain MRI, electroencephalogram, and measures of neurocognitive function and psychiatric symptoms. These assessments include higher level measurement of all cognitive domains (thinking, language processing, memory, attention, and executive functioning) [60], visual motor integration and speed [61–63], and a psychiatric symptom battery [64].

*Biospecimens* are collected across all Tiers using kits designed specifically for each visit, timepoint, and participant age (**Table 5; Supplemental Table 6**). Tier 1 biospecimens consist of saliva and whole blood. Kits are shipped to homes for remote collection. Child and primary caregivers provide both saliva and blood; the other biological parent when available provides only saliva. Saliva is collected using Oragene devices (OGR-600) and banked for future DNA analysis. Whole blood is collected using a TASSO M20 device [65], which collects capillary blood using 4 volumetric sponges that each hold 17.5µL of blood (70 µL total). One sponge is used for SARS-CoV-2 spike and nucleocapsid antibody testing and remaining sponges are banked for future use.

**Table 5:**
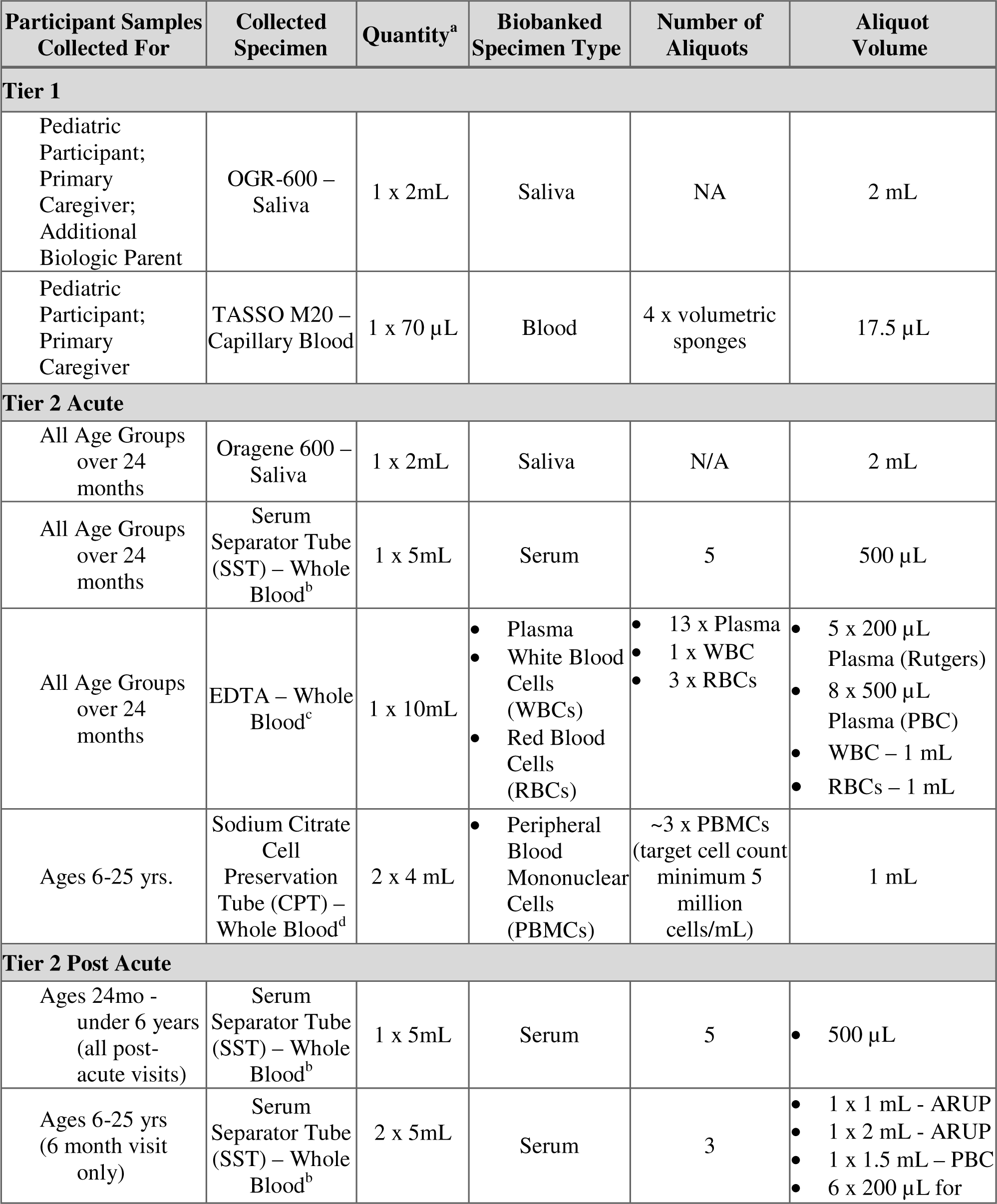
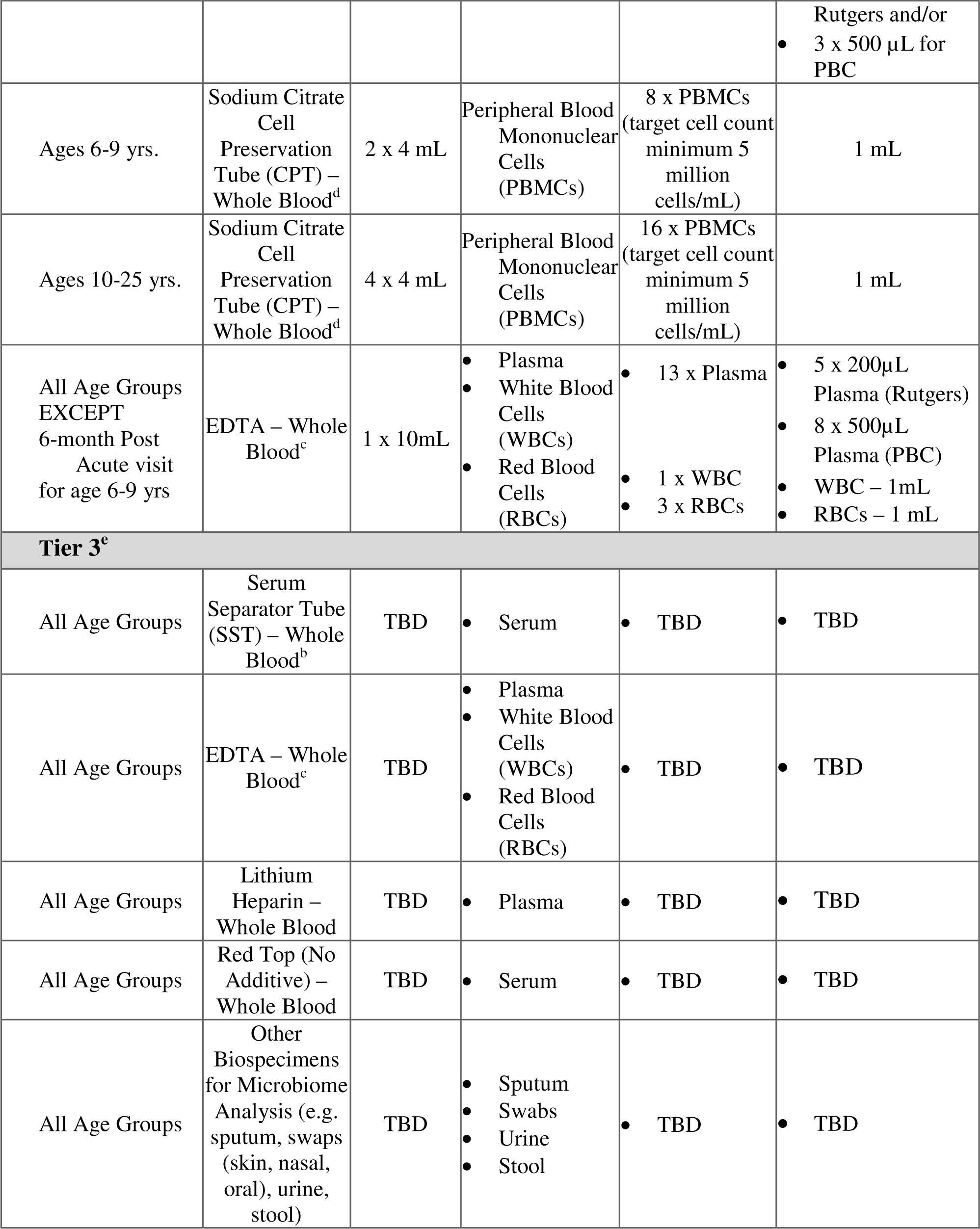
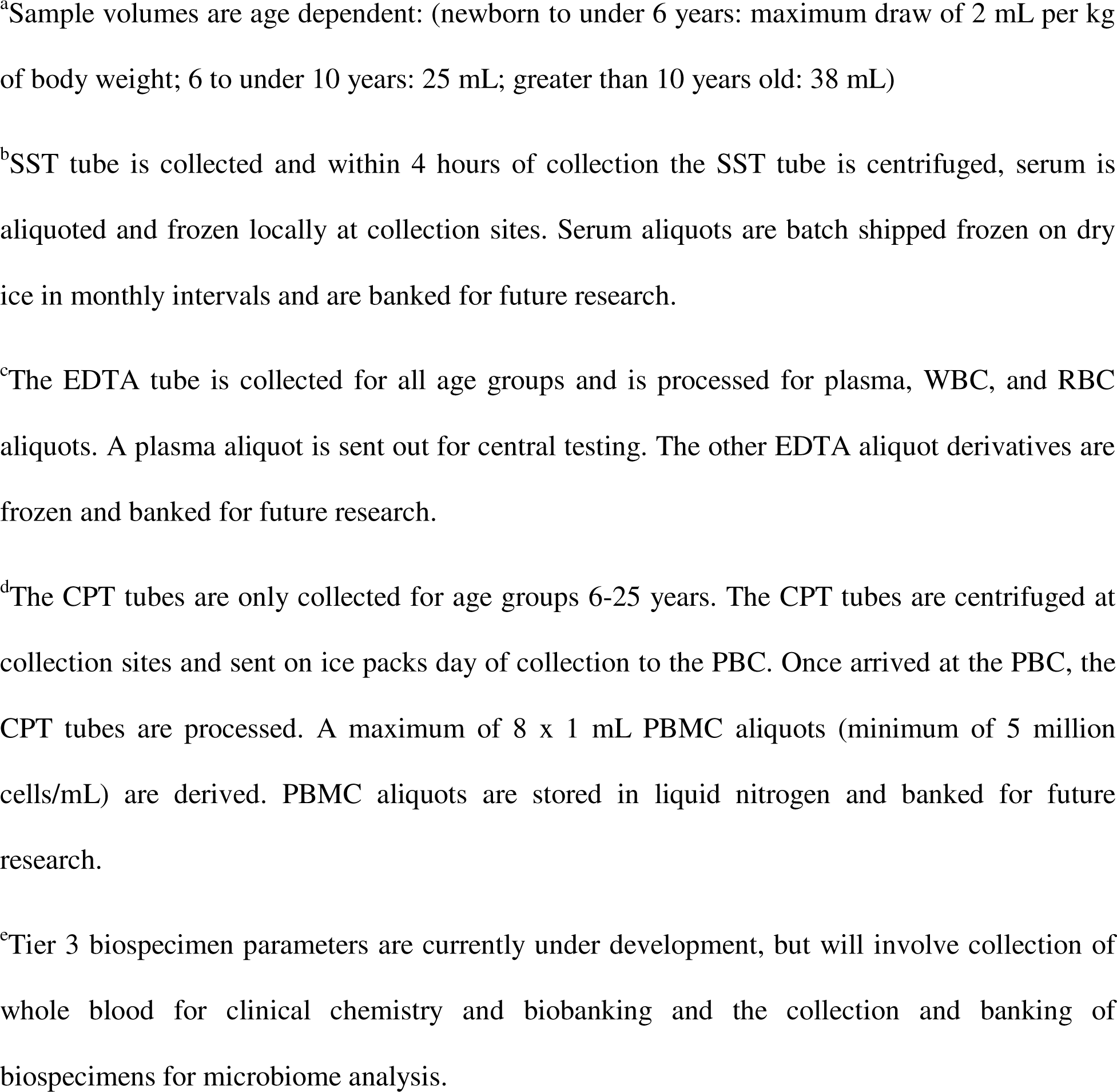
Biospecimen Collection and Processing Summary.

| Participant Samples Collected For | Collected Specimen | Quantity<sup>a</sup> | Biobanked Specimen Type | Number of Aliquots | Aliquot Volume |
| --- | --- | --- | --- | --- | --- |
| **Tier 1** |  |  |  |  |  |
| Pediatric Participant;<br>Primary Caregiver;<br>Additional Biologic Parent | OGR-600 – Saliva | 1 x 2mL | Saliva | NA | 2 mL |
| Pediatric Participant;<br>Primary Caregiver | TASSO M20 – Capillary Blood | 1 x 70 µL | Blood | 4 x volumetric sponges | 17.5 µL |
| **Tier 2 Acute** |  |  |  |  |  |
| All Age Groups over 24 months | Oragene 600 – Saliva | 1 x 2mL | Saliva | N/A | 2 mL |
| All Age Groups over 24 months | Serum Separator Tube (SST) – Whole Blood<sup>b</sup> | 1 x 5mL | Serum | 5 | 500 µL |
| All Age Groups over 24 months | EDTA – Whole Blood<sup>c</sup> | 1 x 10mL | • Plasma<br>• White Blood Cells (WBCs)<br>• Red Blood Cells (RBCs) | • 13 x Plasma<br>• 1 x WBC<br>• 3 x RBCs | • 5 x 200 µL Plasma (Rutgers)<br>• 8 x 500 µL Plasma (PBC)<br>• WBC – 1 mL<br>• RBCs – 1 mL |
| Ages 6-25 yrs. | Sodium Citrate Cell Preservation Tube (CPT) – Whole Blood<sup>d</sup> | 2 x 4 mL | • Peripheral Blood Mononuclear Cells (PBMCs) | ~3 x PBMCs (target cell count minimum 5 million cells/mL) | 1 mL |
| **Tier 2 Post Acute** |  |  |  |  |  |
| Ages 24mo - under 6 years (all post-acute visits) | Serum Separator Tube (SST) – Whole Blood<sup>b</sup> | 1 x 5mL | Serum | 5 | • 500 µL |
| Ages 6-25 yrs (6 month visit only) | Serum Separator Tube (SST) – Whole Blood<sup>b</sup> | 2 x 5mL | Serum | 3 | • 1 x 1 mL - ARUP<br>• 1 x 2 mL - ARUP<br>• 1 x 1.5 mL – PBC<br>• 6 x 200 µL for |
|  |  |  |  |  | Rutgers and/or 3 x 500 µL for PBC |
| Ages 6-9 yrs. | Sodium Citrate Cell Preservation Tube (CPT) – Whole Blood<sup>d</sup> | 2 x 4 mL | Peripheral Blood Mononuclear Cells (PBMCs) | 8 x PBMCs (target cell count minimum 5 million cells/mL) | 1 mL |
| Ages 10-25 yrs. | Sodium Citrate Cell Preservation Tube (CPT) – Whole Blood<sup>d</sup> | 4 x 4 mL | Peripheral Blood Mononuclear Cells (PBMCs) | 16 x PBMCs (target cell count minimum 5 million cells/mL) | 1 mL |
| All Age Groups EXCEPT 6-month Post Acute visit for age 6-9 yrs | EDTA – Whole Blood<sup>c</sup> | 1 x 10mL | Plasma<br>White Blood Cells (WBCs)<br>Red Blood Cells (RBCs) | 13 x Plasma<br>1 x WBC<br>3 x RBCs | 5 x 200µL Plasma (Rutgers)<br>8 x 500µL Plasma (PBC)<br>WBC – 1mL<br>RBCs – 1 mL |
| **Tier 3<sup>e</sup>** |  |  |  |  |  |
| All Age Groups | Serum Separator Tube (SST) – Whole Blood<sup>b</sup> | TBD | Serum | TBD | TBD |
| All Age Groups | EDTA – Whole Blood<sup>c</sup> | TBD | Plasma<br>White Blood Cells (WBCs)<br>Red Blood Cells (RBCs) | TBD | TBD |
| All Age Groups | Lithium Heparin – Whole Blood | TBD | Plasma | TBD | TBD |
| All Age Groups | Red Top (No Additive) – Whole Blood | TBD | Serum | TBD | TBD |
| All Age Groups | Other Biospecimens for Microbiome Analysis (e.g. sputum, swaps (skin, nasal, oral), urine, stool) | TBD | Sputum<br>Swabs<br>Urine<br>Stool | TBD | TBD |

<sup>a</sup>Sample volumes are age dependent: (newborn to under 6 years: maximum draw of 2 mL per kg of body weight; 6 to under 10 years: 25 mL; greater than 10 years old: 38 mL)
<sup>b</sup>SST tube is collected and within 4 hours of collection the SST tube is centrifuged, serum is aliquoted and frozen locally at collection sites. Serum aliquots are batch shipped frozen on dry ice in monthly intervals and are banked for future research.
<sup>c</sup>The EDTA tube is collected for all age groups and is processed for plasma, WBC, and RBC aliquots. A plasma aliquot is sent out for central testing. The other EDTA aliquot derivatives are frozen and banked for future research.
<sup>d</sup>The CPT tubes are only collected for age groups 6-25 years. The CPT tubes are centrifuged at collection sites and sent on ice packs day of collection to the PBC. Once arrived at the PBC, the CPT tubes are processed. A maximum of 8 x 1 mL PBMC aliquots (minimum of 5 million cells/mL) are derived. PBMC aliquots are stored in liquid nitrogen and banked for future research.
<sup>e</sup>Tier 3 biospecimen parameters are currently under development, but will involve collection of whole blood for clinical chemistry and biobanking and the collection and banking of biospecimens for microbiome analysis.

Tier 2 acute biospecimens include saliva (Oragene OGR-600) and whole blood collections. All post-acute Tier 2 biospecimens consist of whole blood. The maximum amount of blood drawn at a single visit is age dependent. Whole blood is collected using serum separator tube (SST) and Ethylenediaminetetraacetic tube acid (EDTA) across all ages above 24 months and an additional cell preparation tube (CPT) is included at participants 6 years of age and older.

Tier 3 biospecimens consist of whole blood, sputum, swabs (e.g., skin, nasal, oral), urine and stool. Collection of Tier 3 biospecimens is limited to children ages 3 years and older (maximum allowable volume is age dependent).

### Statistical methods

We will estimate the proportion of children and young adults experiencing new onset or worsening of each symptom (incidence), stratified by age (0-2, 3-5, 6-12, 13-17, 18-25 years), over time. Prevalence within the recruited population will be estimated by calculating the point prevalence of each symptom at each 3-month interval since infection by calculating the proportion of children and young adults who are currently experiencing each symptom at each study visit. The excess burden of each symptom due to infection will be assessed by calculating differences in incidence and prevalence between children and young adults with and without an infection history. Odds ratios and relative risks for the association between infection and onset of each symptom will also be calculated, adjusting for sex in each age strata.

A preliminary case definition of PASC will be informed by using variable selection methods to identify which symptoms best differentiate children and young adults with and without an infection. The estimated associations obtained from regression models will be used to define a PASC score, with a cutoff for PASC defined based on clinical expertise while ensuring that the rate of those with no history of infection who are diagnosed as having PASC is reasonably low. This preliminary symptom-based case definition will be modified and augmented by clinical and subclinical findings as they become available. To identify PASC phenotypes among children and young adults who are classified as having PASC defined by symptom patterns, we will use unsupervised learning methods to discover symptom clusters within each age strata (e.g., agglomerative hierarchical clustering [66] and consensus clustering [67]) to define PASC sub-phenotypes.

With this definition of PASC, we will conduct regression analyses to evaluate whether the risk of PASC and PASC sub-types differs by multiple factors, including demographic, clinical, and caregiver characteristics, social determinants of health, SARS-CoV-2 infection and immunization history, symptom severity during the acute phase of SARS-CoV-2 infection, and therapeutic exposures. Among participants in Tier 2 who develop PASC, we will use time-to-event analyses to identify factors that influence time to recovery from PASC. To investigate biomarkers related to PASC, clinical laboratory assessments will be compared between children and young adults who do and do not develop PASC. Mediation analyses will also be used to study the pathways by which SARS-CoV-2 infection leads to the development of PASC.

### Power calculations

Power calculations for the *de novo cohort* were performed prior to recruitment using a type 1 error rate of 0.01. With 4,800 infected and 1,200 uninfected children and young adults from both acute and post-acute arms in Tier 1, assuming the risk of a given symptom in the uninfected group is 10%, we have 90% power to detect a difference as small as 4.1% in the frequency of that symptom between groups.

In Tier 2, given the sampling and promotion framework described in *Timing of Study Assessments*, our sample with longitudinal follow-up will be skewed towards those who are more likely to have PASC. Following development of a definition of PASC, we consider the scenario in which we assume that of the 5,400 children and young adults with a history of infection in Tier 2, 3,600 meet PASC criteria and 1,800 do not. For a hypothetical risk factor with 50% prevalence in the PASC-group, we have 90% power to detect an odds ratio as small as 1.25 for the odds of PASC for those with the risk factor versus those without. For a factor with 25% prevalence in the PASC-negative group, the minimum detectable odds ratio is 1.29. In our Tier 3 sample of 600 children and young adults with history of infection (which includes additional data on biomarkers), assuming the sample has 400 with PASC and 200 without PASC, for a marker with 10% prevalence in the PASC-group, we have 90% power to detect an odds ratio as small as 2.60 for PASC.

## Discussion

The overall goal of RECOVER-Pediatrics is to improve our understanding of recovery after SARS-CoV-2 infection, with a focus on the prevalence, natural history, and pathogenesis of PASC in children and young adults. Successful completion should lead to formal characterization of pediatric PASC as its own syndrome. This is essential to develop diagnostic, treatment, and preventive strategies tailored to children’s unique physiology.

RECOVER-Pediatrics is well positioned to ascertain the epidemiology, four-year clinical course, and sociodemographic contributions to pediatric PASC, with rich data and biosamples available to readily test further mechanistic hypotheses, establish biomarkers, and provide insights into potential therapies. The meta-cohort is designed to provide details that are not available in other large epidemiologic or electronic health records queries, including a dynamic study design that can be flexible and responsive as new variants arise, and as our understanding of the long-term effects of SARS-CoV-2 evolves. RECOVER-Pediatrics was designed to include a wide range of ages, and diverse socioeconomic, racial, ethnic and geographic populations to ensure that findings are generalizable, and provide equitable benefit for all.

The generation-defining nature of the COVID-19 pandemic will impact the life course of children in ways that we have yet to fully understand. The unprecedented scope of RECOVER-Pediatrics sets the stage for not only characterizing a new disorder that will impact children for years to come, but also for identifying and deploying solutions through its collaborations with investigators and communities across the country.

RECOVER-Pediatrics is expected to gather a rich data set that can be used to develop treatments for persons with Long COVID and provide guidelines for how to respond more quickly to prevent, reduce the consequences, and treat complications of future coronavirus outbreaks which are likely to emerge.

## Data Availability

No datasets were generated or analysed during the current study. All relevant data from this study will be made available upon study completion.

## Funding

This research was funded by the National Institutes of Health (NIH) Agreement OTA OT2HL161847 as part of the Researching COVID to Enhance Recovery (RECOVER) research Initiative.

## Competing interests

Brett Anderson reported receiving direct support for work not related to RECOVER work/publications from Genentech and the National Institute of Allergy and Immunology.

Walter Dehority reported receiving grant support from Merck and participating in research for the Moderna COVID-19 pediatric vaccine trial and the Pfizer Paxlovid trial.

Alex Fiks reported receiving support from NJM insurance and personal consulting fees not related to this paper from Rutgers University and the American Academy of Pediatrics.

Ashraf Harahsheh reported serving as a scientific advisory board member unrelated to this paper for OP2 DRUGS.

Lawrence Kleinman reported serving as an unpaid member of the Board of Directors for the DARTNet Institute, as a principle investigator at Quality Matters, Inc., and as the Vice Chair for the Borough of Metuchen Board of Health. Dr. Kleinman also reported grant support for work not related to RECOVER work/publications from NIH, HRSA, and the Robert Wood Johnson Foundation. Dr. Kleinman also reported minority individual stock ownership in Apple Computer, Sanofi SA, Experion, GlaxoSmithKline, Magyar Bank, Regeneron Pharmaceuticals, JP Morgan Chase, and Amgen Inc.

Torri Metz reported participating as a Principle Investigator in the medical advisory board for the planning of a Pfizer clinical trial of SARS-CoV-2 vaccination in pregnancy. She is also a principle investigator for a Pfizer study evaluating the pharmacokinetics of Paxlovid in pregnant people with COVID-19.

Joshua Milner reported serving as a member of the Scientific Advisory Board for Blueprint Medicines, in a capacity unrelated to RECOVER work/publications.

## Disclaimer

Authorship has been determined according to ICMJE recommendations. The content is solely the responsibility of the authors and does not necessarily represent the official views of the RECOVER program, the NIH or other funders.

## Acknowledgement

We would like to thank the National Community Engagement Group (NCEG), all patient, caregiver and community representatives, and all the participants enrolled in the RECOVER initiative.

## Supporting information

The supporting information is provided in a supplemental appendix file.

## Notes

### Competing Interest Statement

I have read the journal's policy and the authors of this manuscript have the following competing interests: Brett Anderson reported receiving direct support for work not related to RECOVER work/publications from Genentech and the National Institute of Allergy and Immunology. Walter Dehority reported receiving grant support from Merck and participating in research for the Moderna COVID-19 pediatric vaccine trial and the Pfizer Paxlovid trial. Alex Fiks reported receiving support from NJM insurance and personal consulting fees not related to this paper from Rutgers University and the American Academy of Pediatrics. Ashraf Harahsheh reported serving as a scientific advisory board member unrelated to this paper for OP2 DRUGS. Lawrence Kleinman reported serving as an unpaid member of the Board of Directors for the DARTNet Institute, as a principle investigator at Quality Matters, Inc., and as the Vice Chair for the Borough of Metuchen Board of Health. Dr. Kleinman also reported grant support for work not related to RECOVER work/publications from NIH, HRSA, and the Robert Wood Johnson Foundation. Dr. Kleinman also reported minority individual stock ownership in Apple Computer, Sanofi SA, Experion, GlaxoSmithKline, Magyar Bank, Regeneron Pharmaceuticals, JP Morgan Chase, and Amgen Inc. Torri Metz reported participating as a Principle Investigator in the medical advisory board for the planning of a Pfizer clinical trial of SARS-CoV-2 vaccination in pregnancy. She is also a principle investigator for a Pfizer study evaluating the pharmacokinetics of Paxlovid in pregnant people with COVID-19. Joshua Milner reported serving as a member of the Scientific Advisory Board for Blueprint Medicines, in a capacity unrelated to RECOVER work/publications.

### Clinical Protocols

https://recovercovid.org/sites/default/files/docs/PediatricProtocol.v2.3.pdf

### Funding Statement

National Institutes of Health (NIH) Agreement OTA OT2HL161847 (SDK, RG), OT2HL161841 (ASF). https://www.nih.gov/ The funders did not and will not have a role in study design, data collection and analysis, decision to publish, or preparation of the manuscript.

### Author Declarations

IRB of NYU Grossman School of Medicine gave ethical approval for this work.

### Summary of Updates

We have updated the manuscript file to reflect equal first author contributions and group authorship on behalf of the RECOVER Initiative, per the PLOS One formatting guidelines.

