## Supplemental Appendix for "Researching COVID to enhance recovery (RECOVER) pediatric study protocol: Rationale, objectives and design"

### Supplemental Tables

#### Supplemental Table 1: Hubs and enrolling sites

| Cohort Type | Hub Name | Enrolling Sites | Location |
| --- | --- | --- | --- |
| *de novo* pediatric RECOVER cohort | Arkansas Children’s Research Institute | Arkansas Children’s Research Institute | Arkansas |
|  | Arkansas Children’s Research Institute | Avera Research Institute | South Dakota |
|  | Arkansas Children’s Research Institute | Dartmouth Hitchcock Medical Center | New Hampshire |
|  | Arkansas Children’s Research Institute | Kapiolani Medical Center for Women and Children | Hawaii |
|  | Arkansas Children’s Research Institute | Medical University of South Carolina | South Carolina |
|  | Arkansas Children’s Research Institute | Nemours Children’s Health System | Delaware |
|  | Arkansas Children’s Research Institute | Northeastern University, Puerto Rico Testsite | Puerto Rico |
|  | Arkansas Children’s Research Institute | Pennington Biomedical Research Center | Louisiana |
|  | Arkansas Children’s Research Institute | University of Louisville Research Foundation | Kentucky |
|  | Arkansas Children’s Research Institute | University of Nebraska Medical Center | Nebraska |
|  | Arkansas Children’s Research Institute | University of New Mexico Health Sciences Center | New Mexico |
|  | Arkansas Children’s Research Institute | University of Oklahoma Health Sciences Center | Oklahoma |
|  | Arkansas Children’s Research Institute | University of Vermont Medical Center | Vermont |
|  | Arkansas Children’s Research Institute | West Virginia University | West Virginia |
|  | Children’s Hospital of Los Angeles | Children’s Hospital of Los Angeles | California |
|  | Columbia University College of Physicians & Surgeons | Columbia University College of Physicians & Surgeons | New York |
|  | Columbia University College of Physicians & Surgeons | Best Healthcare Inc. | New York |
|  | Rutgers Robert Wood Johnson Medical Center | Rutgers Robert Wood Johnson Medical Center | New Jersey |
|  | Rutgers Robert Wood Johnson Medical Center | American Academy of Family Physicians, AAFP, NRN, | National reach |
|  | Rutgers Robert Wood Johnson Medical Center | American Academy of Pediatrics | National reach |
|  | Rutgers Robert Wood Johnson Medical Center | Children's Mercy Kansas City | Missouri |
|  | Rutgers Robert Wood Johnson Medical Center | Connecticut Children's Medical Center | Connecticut |
|  | Rutgers Robert Wood Johnson Medical Center | Hackensack Meridian Health Hospitals Corporation | New Jersey |
|  | Rutgers Robert Wood Johnson Medical Center | The MetroHealth System | Ohio |
|  | Rutgers Robert Wood Johnson Medical Center | New York Medical Center, Westchester Medical Center | New York |
|  | Rutgers Robert Wood Johnson Medical Center | Saint Barnabas Medical Center | New Jersey |
|  | Rutgers Robert Wood Johnson Medical Center | Yale School of Medicine | Connecticut |
|  | University of California San Diego/Rady Children’s Hospital | University of California San Diego/Rady Children’s Hospital | California |
|  | Virginia Commonwealth University | Virginia Commonwealth University | Virginia |
|  | Virginia Commonwealth University | Rhode Island Hospital | Rhode Island |
|  | Virginia Commonwealth University | NYU Langone Health | New York |
| Adolescent Brain Cognitive Development (ABCD) | University of California San Diego | University of California San Diego | California |
|  | University of California San Diego | Children's Hospital, Los Angeles | California |
|  | University of California San Diego | Florida International University | Florida |
|  | University of California San Diego | Laureate Institute for Brain Research | Oklahoma |
|  | University of California San Diego | Medical University of South Carolina | South Carolina |
|  | University of California San Diego | Oregon Health & Science University | Oregon |
|  | University of California San Diego | SRI International | California |
|  | University of California San Diego | University of California, Los Angeles | California |
|  | University of California San Diego | University of Colorado Boulder | Colorado |
|  | University of California San Diego | University of Florida | Florida |
|  | University of California San Diego | University of Maryland Baltimore | Maryland |
|  | University of California San Diego | University of Michigan | Michigan |
|  | University of California San Diego | University of Minnesota | Minnesota |
|  | University of California San Diego | University of Pittsburgh Medical Center | Pennsylvania |
|  | University of California San Diego | University of Rochester | New York |
|  | University of California San Diego | University of Utah | Utah |
|  | University of California San Diego | University of Vermont | Vermont |
|  | University of California San Diego | University of Wisconsin, Milwaukee | Wisconsin |
|  | University of California San Diego | Virginia Commonwealth University | Virginia |
|  | University of California San Diego | Washington University St. Louis | Missouri |
|  | University of California San Diego | Yale University | Connecticut |
| COVID MUSIC Study | HealthCore, Inc | Ann & Robert Lurie Children's Hosp | Illinois |
|  | HealthCore, Inc | Baylor/Texas Children’s Hospital | Texas |
|  | HealthCore, Inc | Boston Children’s Hospital | Massachusetts |
|  | HealthCore, Inc | Children’s Healthcare of Atlanta | Georgia |
|  | HealthCore, Inc | Children’s Hospital of Colorado | Colorado |
|  | HealthCore, Inc | Children’s Hospital Los Angeles | California |
|  | HealthCore, Inc | Children’s Hospital of Michigan | Michigan |
|  | HealthCore, Inc | Children’s Hospital of New Orleans | Louisiana |
|  | HealthCore, Inc | Children's Hospital of Philadelphia (CHOP) | Pennsylvania |
|  | HealthCore, Inc | Children’s Mercy Hospital | Missouri |
|  | HealthCore, Inc | Children's National Hospital | Washington DC |
|  | HealthCore, Inc | Cincinnati Children’s Hospital Medical Center | Ohio |
|  | HealthCore, Inc | Cohen Children’s Medical Center | New York |
|  | HealthCore, Inc | CS Mott Children’s Hospital/University of Michigan | Michigan |
|  | HealthCore, Inc | Dell Children’s Medical Center | Texas |
|  | HealthCore, Inc | Hospital for Sick Children, Toronto | Toronto |
|  | HealthCore, Inc | Joe DiMaggio Children’s Hospital | Florida |
|  | HealthCore, Inc | Medical College of Wisconsin, Children's Hospital | Wisconsin |
|  | HealthCore, Inc | Medical University of South Carolina | South Carolina |
|  | HealthCore, Inc | Morgan Stanley Children's Hospital | New York |
|  | HealthCore, Inc | Nemours, Alfred I. duPont Hospital for Children | Delaware |
|  | HealthCore, Inc | Phoenix Children’s Hospital | Arizona |
|  | HealthCore, Inc | Primary Children’s Hospital/University of Utah | Utah |
|  | HealthCore, Inc | Rady Children’s Hospital | California |
|  | HealthCore, Inc | Riley Children’s Hospital | Indiana |
|  | HealthCore, Inc | Seattle Children’s Hospital | Washington |
|  | HealthCore, Inc | University of Alabama | Alabama |
|  | HealthCore, Inc | University of Mississippi | Mississippi |
|  | HealthCore, Inc | UT Southwestern, Children's Health Dallas | Texas |
|  | HealthCore, Inc | Valley Children's Healthcare and Hospital | California |
| Post-COVID vaccine myocarditis cohort | HealthCore, Inc | HealthCore, Inc | Massachusetts |
| In-utero exposure cohort | University of California San Francisco | University of California San Francisco, | California and nationally through home visits |
|  | NICHD Maternal-Fetal Medicine Units (MFMU) Network | University of Utah | Utah |
|  | NICHD Maternal-Fetal Medicine Units (MFMU) Network | Brown University Women and Infants Hospital | Rhode Island |
|  | NICHD Maternal-Fetal Medicine Units (MFMU) Network | Case Western MetroHealth Medical | Ohio |
|  | NICHD Maternal-Fetal Medicine Units (MFMU) Network | ChristianaCare Health | Delaware |
|  | NICHD Maternal-Fetal Medicine Units (MFMU) Network | Columbia University | New York |
|  | NICHD Maternal-Fetal Medicine Units (MFMU) Network | Duke University Medical Center | North Carolina |
|  | NICHD Maternal-Fetal Medicine Units (MFMU) Network | Good Samaritan | New York |
|  | NICHD Maternal-Fetal Medicine Units (MFMU) Network | Miami Valley Hospital | Ohio |
|  | NICHD Maternal-Fetal Medicine Units (MFMU) Network | New York-Presbyterian, Queens | New York |
|  | NICHD Maternal-Fetal Medicine Units (MFMU) Network | NorthShore University HealthSystem | Illinois |
|  | NICHD Maternal-Fetal Medicine Units (MFMU) Network | Northwestern University | Illinois |
|  | NICHD Maternal-Fetal Medicine Units (MFMU) Network | Ohio State University | Ohio |
|  | NICHD Maternal-Fetal Medicine Units (MFMU) Network | Saint Peter's University Hospital | New Jersey |
|  | NICHD Maternal-Fetal Medicine Units (MFMU) Network | UH MacDonald's Women's Hospital | Ohio |
|  | NICHD Maternal-Fetal Medicine Units (MFMU) Network | University of Alabama at Birmingham | Alabama |
|  | NICHD Maternal-Fetal Medicine Units (MFMU) Network | University of North Carolina, Chapel Hill | North Carolina |
|  | NICHD Maternal-Fetal Medicine Units (MFMU) Network | University of Pennsylvania | Pennsylvania |
|  | NICHD Maternal-Fetal Medicine Units (MFMU) Network | University of Pittsburgh | Pennsylvania |
|  | NICHD Maternal-Fetal Medicine Units (MFMU) Network | University of Texas HSC at Houston - Memorial City | Texas |
|  | NICHD Maternal-Fetal Medicine Units (MFMU) Network | University of Texas HSC at Houston, LBJ Hospital | Texas |
|  | NICHD Maternal-Fetal Medicine Units (MFMU) Network | University of Texas HSC at Houston, Memorial Herma | Texas |
|  | NICHD Maternal-Fetal Medicine Units (MFMU) Network | University of Texas Medical Branch at Galveston | Texas |
|  | NICHD Maternal-Fetal Medicine Units (MFMU) Network | Wakemed Raleigh/Wakemed North | North Carolina |
|  | NICHD Maternal-Fetal Medicine Units (MFMU) Network | Yale University | Connecticut |

#### Supplemental Table 2. Inclusion and exclusion criteria

| **Inclusion Criteria** | **Exclusion Criteria** |
| --- | --- |
| - Birth through 25 years old - Any SARS-CoV-2 infection status (never infected, suspected, probable, or confirmed) | - Co-morbid illness with expected survival less than 2 years - Any child or young adult who in the opinion of the site investigator may be at increased risk of adverse events during participation in the study, or who may not be able to complete study procedures due to co-morbid disease or disability - Any young adult above the age of majority who lacks capacity to provide consent - Any child with a plan for adoption or where the state is the legal guardian - Any young adult who is incarcerated, or who lacks capacity to provide consent - Young adult currently enrolled in RECOVER-Adult or RECOVER-Pregnancy |

Note: Participants are eligible without exclusion related to sex, race/ethnicity, geography, nationality, severity of disease, underlying health conditions, the presense or absence of PASC symptoms, or COVID vaccine status.

#### Supplemental Table 3: Inclusion into analytic groups.

**“Infected”: Children and Young Adults with a History of SARS-CoV-2 Infection**

- Ages newborn through 25 years old
- Suspected, probable, or confirmed SARS-CoV-2 infection as defined by WHO criteria since January 1, 2020
- Children/young adults with or without history of MIS-C
- Children/young adults with or without history of SARS-CoV-2 vaccination
- Children/young adults with evidence of past SARS-CoV-2 infection based on serum antibody profile (with or without history of acute symptoms)
- Children/young adults with recurrent SARS-CoV-2 infections and those with post-vaccination (breakthrough) infections

**“Uninfected”: Children and Young Adults without a Known History of SARS-CoV-2 Infection**

- Does not meet WHO criteria for a suspected, probable, or confirmed case of SARS-CoV-2 infection AND
- Does not have serological evidence of past asymptomatic SARS-CoV-2 infection in medical history or Tier 1 testing, AND
- Lives in the same communities or recruited from the same sources as those in the SARS-CoV-2 infected cohort, AND
- Either not hospitalized for any reason in prior 3 months, or hospitalized (with or without ICU stay) within the prior 3 months

Note: Uninfected individuals who develop SARS-CoV-2 infection during the study period will be reassigned to the SARS-Cov-2 infected group and will be considered to have been enrolled prior to SARS-CoV-2 infection.

#### Supplemental Table 4: World Health Organization (WHO) criteria.

**Children and Young Adults with Suspected SARS-Cov-2 Infection**

a. Children/young adults who meet these clinical criteria:

At least one of these clinical criteria:

- Acute onset of fever and cough OR
- Acute onset of any three or more of the following signs or symptoms: fever, cough, general weakness /fatigue, headache, myalgia, sore throat, coryza, dyspnea, anorexia/nausea/vomiting, diarrhea, altered mental status.

AND at least one of these epidemiological criteria:

- Residing or working in an area with a high risk of transmission of virus: closed residential, school or camp settings anytime within the 14 days before symptom onset; OR
- Residing or travel to an area with community transmission anytime within the 14 days before symptom onset; OR
- Any known household contact or any member of the household working in any health care setting, including within health facilities or within the community; anytime within the 14 days before symptom onset.

b. Patient with history of severe acute respiratory illness (SARI): acute respiratory infection with history of fever or measured fever of ≥ 38 C°; and cough; with onset within the last 10 days; and requires hospitalization
c. An asymptomatic person not meeting epidemiologic criteria with a positive SARS-CoV-2 Antigen-RDT

**Children and Young Adults with Probable SARS-Cov-2 Infection**

a. A patient who meets clinical criteria above AND is a contact of a probable or confirmed case or linked to a COVID-19 cluster; OR
b. A suspect case with chest imaging showing findings suggestive of COVID-19 disease; OR
c. A person with recent onset of anosmia (loss of smell) or ageusia (loss of taste) in the absence of any other identified cause

**Children and Young Adults with Confirmed SARS-Cov-2 Infection**

a. A person with a positive Nucleic Acid Amplification Test (NAAT); OR
b. A person with a positive SARS-CoV-2Antigen-RDT AND meeting either the probable case definition or suspect criteria A OR B; OR
c. An asymptomatic person with a positive SARS-CoV-2 Antigen-RDT who is a contact of a probable or confirmed case

**Children and Young Adults with Asymptomatic SARS-CoV-2 Infection**

a. A person without history of acute COVID-19 symptoms who has one or more of the epidemiological exposures for suspected infection and who also meets criteria b or c for suspected or probable infection, or who meets any of the criteria for confirmed infection
b. A person without history of acute COVID-19 symptoms who has positive nucleocapsid antibody test result in medical history or Tier 1 testing with or without NAAT or RDT testing or known contact to a probable or confirmed case

#### Supplemental Table 5: Survey topics in tiers 1 and 2 questionnaires.

| Survey Instrument | Topic | Asked in Tier 2  follow-up surveys | Source of survey,  if not developed for RECOVER |
| --- | --- | --- | --- |
| Household-Level and Child-Level Surveys | | | |
| Demographics | Name and contact information | ✓ |  |
| Demographics | Alternate contacts | ✓ |  |
| Demographics | Date of birth |  |  |
| Demographics | Sex assigned at birth |  | All of Us Research Program |
| Demographics | Gender identity |  | All of Us Research Program |
| Demographics | Race and ethnicity |  | All of Us Research Program |
| Demographics | Languages spoken |  | California Health Interview Survey |
| Demographics | Country of origin |  | American Community Survey (ACS) |
| Demographics | Educational attainment (grade, school type) |  | National Health and Nutrition Examination Survey (NHANES) |
| Child birth history | Birth mother age |  | National Survey of Children’s Health |
| Child birth history | Child birth weight |  | National Survey of Children’s Health |
| Child birth history | Child birth length |  | National Survey of Children’s Health |
| Child birth history | Prematurity/  gestational age |  | National Survey of Children’s Health |
| Child birth history | Delivery type |  |  |
| Child birth history | NICU admision |  |  |
| Child birth history | Pregnancy complications |  |  |
| Child birth history | Breastfeeding |  |  |
| Child current health status | Child current length  or height | ✓ | National Survey of Children’s Health |
| Child current health status | Child current weight | ✓ | National Survey of Children’s Health |
| Child current health status | Biological parents' height and weight |  |  |
| Child current health status | Child menses | ✓ |  |
| Child current health status | Child disabilities |  | National Survey of Children’s Health |
| Child current health status | Household smoking exposure | ✓ |  |
| Special Health Care Needs Screener | Special Health Care Needs Screener | ✓ | Children with Special Health Care Needs (CSHCN) Screener |
| Special Health Care Needs Screener | Asthma | ✓ | National Survey of Children’s Health |
| Special Health Care Needs Screener | Cerebral Palsy | ✓ | National Survey of Children’s Health |
| Special Health Care Needs Screener | Diabetes | ✓ | National Survey of Children’s Health |
| Special Health Care Needs Screener | Epilepsy or seizure disorder | ✓ | National Survey of Children’s Health |
| Special Health Care Needs Screener | Heart problem | ✓ | National Survey of Children’s Health |
| Special Health Care Needs Screener | Frequent or severe headaches, including migraines | ✓ | National Survey of Children’s Health |
| Special Health Care Needs Screener | Tourette's syndrome or tics | ✓ | National Survey of Children’s Health |
| Special Health Care Needs Screener | Anxiety (feeling nervous or anxious) | ✓ | National Survey of Children’s Health |
| Special Health Care Needs Screener | Depression (feeling very sad) | ✓ | National Survey of Children’s Health |
| Special Health Care Needs Screener | Down syndrome | ✓ | National Survey of Children’s Health |
| Special Health Care Needs Screener | Blood disorders | ✓ | National Survey of Children’s Health |
| Special Health Care Needs Screener | Cystic fibrosis | ✓ | National Survey of Children’s Health |
| Special Health Care Needs Screener | Other genetic or inherited condition | ✓ | National Survey of Children’s Health |
| Special Health Care Needs Screener | Problems with behavior | ✓ | National Survey of Children’s Health |
| Special Health Care Needs Screener | Developmental delay | ✓ | National Survey of Children’s Health |
| Special Health Care Needs Screener | Intellectual disability | ✓ | National Survey of Children’s Health |
| Special Health Care Needs Screener | Speech or other language disorder (problems with talking or understanding words) | ✓ | National Survey of Children’s Health |
| Special Health Care Needs Screener | Learning disability (problem with learning) | ✓ | National Survey of Children’s Health |
| Special Health Care Needs Screener | Autism or Autism Spectrum Disorder (ASD) | ✓ | National Survey of Children’s Health |
| Special Health Care Needs Screener | Attention Deficit Disorder (ADD) or Attention Deficit/Hyperactivity Disorder (ADHD) | ✓ | National Survey of Children’s Health |
| Special Health Care Needs Screener | Eating disorders (like Anorexia or Binge eating disorder) | ✓ | National Survey of Children’s Health |
| Special Health Care Needs Screener | Other health problems | ✓ |  |
| Global Health | Self-reported or caregiver-reported overall, physical and mental health | ✓ | Early Childhood Parent Report Global Health 8a; PROMIS Parent Proxy Scale v1.0 – Global Health 7; PROMIS-10 v1.2 |
| COVID infection history | Infection date | ✓ |  |
| COVID infection history | How family learned about COVID infection | ✓ |  |
| COVID infection history | Presence of symptoms | ✓ |  |
| COVID infection history | Duration of symptoms | ✓ |  |
| COVID infection history | Symptom severity | ✓ |  |
| COVID infection history | Health care utilization during COVID infection | ✓ |  |
| COVID infection history | COVID treatments | ✓ |  |
| Related conditions | Multisystem Inflammatory Syndrome in Children (MIS-C) | ✓ |  |
| Related conditions | POTS (Postural Orthostatic Tachycardia Syndrome) or other form of dysautonomia or autonomic dysfunction | ✓ |  |
| Related conditions | Long COVID diagnosis | ✓ |  |
| COVID Testing History | Testing history |  |  |
| COVID Testing History | Testing access |  |  |
| COVID Family Infection | COVID infection |  |  |
| COVID Family Infection | COVID-related hospitalization |  |  |
| COVID Family Infection | COVID-related death | ✓ |  |
| COVID Symptoms | General symptoms or problems | ✓ |  |
| COVID Symptoms | Symptoms or problems in the eyes, ears, nose, and throat | ✓ |  |
| COVID Symptoms | Symptoms or problems involving the heart and lungs | ✓ |  |
| COVID Symptoms | Symptoms or problems involving the belly | ✓ |  |
| COVID Symptoms | Symptoms or problems involving the skin, hair, and nails | ✓ |  |
| COVID Symptoms | Symptoms or problems involving the bones and muscles | ✓ |  |
| COVID Symptoms | Symptoms or problems involving the brain and nerves | ✓ |  |
| COVID Symptoms | Symptoms or problems involving feelings or behavior | ✓ |  |
| COVID Symptoms | Symptoms or problems involving periods | ✓ |  |
| COMPASS-31 | Symptoms associated with dysautonomia | ✓ | COMPASS-31 |
| COVID vaccine history | Child COVID vaccine history | ✓ |  |
| COVID vaccine history | Birth mother COVID vaccine history while pregnant |  |  |
| COVID vaccine history | Birth mother COVID vaccine history while breastfeeding |  |  |
| COVID vaccine history | COVID vaccine intentions |  |  |
| COVID Health Consequences | Perceived weight status | ✓ | Youth Risk Behavior Survey |
| COVID Health Consequences | Child diet | ✓ | Youth Risk Behavior Survey |
| COVID Health Consequences | Physical activity | ✓ | Youth Risk Behavior Survey |
| COVID Health Consequences | Outdoor play | ✓ |  |
| COVID Health Consequences | Screen time | ✓ | Youth Risk Behavior Survey |
| COVID Health Consequences | Sleep | ✓ | Youth Risk Behavior Survey |
| COVID Health Consequences | School disruption | ✓ |  |
| COVID Health Consequences | Grades | ✓ |  |
| COVID Health Consequences | Developmental services (Early intervention, Individualized Education Programs, home visiting) | ✓ |  |
| COVID Health Consequences | Discipline | ✓ | Quick Parenting Assessment |
| COVID Health Consequences | Caregiver-child relationship quality | ✓ | Adult Child Relationship Scale |
| COVID Health Consequences | Cognitive stimulation (reading, teaching, playing, talking) | ✓ | StimQ cognitive home environment questionnaire (self-report version of infant/toddler; preschool; elementary school age) |
| Social Determinants Of Health | Household composition |  |  |
| Social Determinants Of Health | Birth order |  |  |
| Social Determinants Of Health | Housing |  |  |
| Social Determinants Of Health | Marital status |  |  |
| Social Determinants Of Health | Health care utilization | ✓ | National Survey of Children’s Health |
| Social Determinants Of Health | Health literacy |  | Brief health literacy screener |
| Social Determinants Of Health | Health insurance | ✓ |  |
| Social Determinants Of Health | Unmet needs | ✓ |  |
| Social Determinants Of Health | Health care access | ✓ |  |
| Social Determinants Of Health | COVID-related guidelines (e.g., masking, social distancing) |  |  |
| Social Determinants Of Health | Financial difficulties | ✓ |  |
| Social Determinants Of Health | Financial assistance programs | ✓ |  |
| Social Determinants Of Health | Food insecurity | ✓ | USDA Core Food Security Module |
| Social Determinants Of Health | Perceived neighborhood safety | ✓ |  |
| Social Determinants Of Health | Neighhood cohesion | ✓ |  |
| Social Determinants Of Health | Discrimination | ✓ | Everyday Discrimination Scale |
| Social Determinants Of Health | Early childhood experiences |  | Other Childhood Stressors |
| Social Determinants Of Health | Mental health | ✓ | DSM-5 Cross-Cutting Symptom Measure |
| Caregiver-Level Surveys | | | |
| Identity | Caregiver relationship to child |  |  |
| Demographics | Caregiver date of birth |  |  |
| Demographics | Caregiver sex assigned at birth |  | All of Us Research Program |
| Demographics | Caregiver gender identity |  | All of Us Research Program |
| Demographics | Caregiver race and ethnicity |  | All of Us Research Program |
| Demographics | Caregiver languages spoken |  | California Health Interview Survey |
| Demographics | Caregiver country of origin |  | American Community Survey (ACS) |
| Global Health | Caregiver overall, physical and mental health | ✓ | PROMIS global health scale |
| Current health status | Caregiver disabilities |  |  |
| COVID infection history | Caregiver infection date | ✓ |  |
| COVID infection history | How caregiver learned about their own COVID infection | ✓ |  |
| COVID infection history | Caregiver presence of symptoms | ✓ |  |
| COVID infection history | Caregiver duration of symptoms | ✓ |  |
| COVID infection history | Caregiver symptom severity | ✓ |  |
| COVID infection history | Caregiver health care utilization during COVID infection | ✓ |  |
| COVID infection history | Caregiver COVID treatments | ✓ |  |
| COVID Testing History | Caregiver testing history |  |  |
| COVID vaccine history | Caregiver COVID vaccine history | ✓ |  |
| COVID Symptoms | Caregiver general symptoms or problems | ✓ |  |
| COVID Symptoms | Caregiver symptoms or problems in the eyes, ears, nose, and throat | ✓ |  |
| COVID Symptoms | Caregiver symptoms or problems involving the heart and lungs | ✓ |  |
| COVID Symptoms | Symptoms or problems involving the belly | ✓ |  |
| COVID Symptoms | Caregiver symptoms or problems involving the skin, hair, and nails | ✓ |  |
| COVID Symptoms | Caregiver symptoms or problems involving the bones and muscles | ✓ |  |
| COVID Symptoms | Caregiver symptoms or problems involving the brain and nerves | ✓ |  |
| COVID Symptoms | Caregiver symptoms or problems involving feelings or behavior | ✓ |  |
| COVID Symptoms | Caregiver symptoms or problems involving periods | ✓ |  |
| COVID Health Consequences | Caregiver perceived weight status | ✓ | Behavioral Risk Factor Surveillance System (BRFSS) |
| COVID Health Consequences | Caregiver diet | ✓ | Behavioral Risk Factor Surveillance System (BRFSS) |
| COVID Health Consequences | Caregiver physical activity | ✓ | Behavioral Risk Factor Surveillance System (BRFSS) |
| COVID Health Consequences | Caregiver screen time | ✓ | Behavioral Risk Factor Surveillance System (BRFSS) |
| COVID Health Consequences | Caregiver sleep | ✓ | Behavioral Risk Factor Surveillance System (BRFSS) |
| Social Determinants Of Health | Caregiver work | ✓ |  |
| Social Determinants Of Health | Caregiver health insurance | ✓ |  |
| Social Determinants Of Health | Caregiver health care utilization | ✓ |  |
| Social Determinants Of Health | Caregiver positive childhood experiences (PCEs) |  | Positive Childhood Experiences (PCEs) |
| Social Determinants Of Health | Caregiver discrimination | ✓ | Everyday Discrimination Scale |
| Social Determinants Of Health | Caregiver social support | ✓ | RAND Social Support Survey |
| Caregiver wellbeing | Caregiver depressive symptoms | ✓ | Patient Health Questionnaire-9 |
| Caregiver wellbeing | Caregiver anxiety symptoms | ✓ | Generalized Anxiety Disorder-7 |
| Caregiver wellbeing | Caregiver stress | ✓ | Perceived Stress Scale |
| Caregiver wellbeing | Caregiver mental health | ✓ | DSM-5 Cross-Cutting Symptom Measure |

#### Supplemental Table 6: Clinical and laboratory assessments across the tiers in the *de novo* RECOVER-Pediatrics cohort.

| Category | Assessment | Tier 1 | Tier 2 | Tier 3 |
| --- | --- | --- | --- | --- |
| Clinical Assessment | Weight |  | ✓ |  |
| Clinical Assessment | Height or length |  | ✓ |  |
| Clinical Assessment | Waist circumference |  | ✓ |  |
| Clinical Assessment | Skin fold thickness  (triceps and subscapular) |  | ✓ |  |
| Clinical Assessment | Temperature |  | ✓ |  |
| Clinical Assessment | Heart rate |  | ✓ |  |
| Clinical Assessment | Respiratory rate |  | ✓ |  |
| Clinical Assessment | Oxygen saturation |  | ✓ |  |
| Clinical Assessment | Blood pressure |  | ✓ |  |
| Clinical Assessment | 10 Minute Active Standing Test (Assessing blood pressure and heart rate after 5 minutes supine, then after standing for 1, 3, 5, 7 and 10 minutes) |  | ✓ |  |
| Clinical Assessment | Electrocardiogram |  | ✓ |  |
| Clinical Assessment | Spirometry |  | ✓ |  |
| Clinical Assessment | Beighton Scale for Joint Hypermobility |  | ✓ |  |
| Clinical Assessment | NIH toolbox |  | ✓ |  |
| Clinical Assessment | Echocardiogram |  |  | ✓ |
| Clinical Assessment | Cardiac MRI without contrast |  |  | ✓ |
| Clinical Assessment | Pulmonary Function Tests (PFTs) |  |  | ✓ |
| Clinical Assessment | Lung Microbiome (Sputum Induction) |  |  | ✓ |
| Clinical Assessment | Cardiopulmonary Exercise Testing |  |  | ✓ |
| Clinical Assessment | Abdominal ultrasound |  |  | ✓ |
| Clinical Assessment | Brain MRI without contrast |  |  | ✓ |
| Clinical Assessment | Brain EEG |  |  | ✓ |
| Clinical Assessment | Neurocognitive testing |  |  | ✓ |
| Laboratory study | SARS-CoV-2 spike and nucleocapsid antibody | ✓ | ✓ |  |
| Laboratory study | Complete metabolic panel |  | ✓ |  |
| Laboratory study | Complete blood count |  | ✓ |  |
| Laboratory study | Anti nuclear antibody (ANA) |  | ✓ |  |
| Laboratory study | Anti-cyclic citrullinated peptide antibodies (Anti-CCP) |  | ✓ |  |
| Laboratory study | Anti dsDNA antibody |  | ✓ |  |
| Laboratory study | Rheumatoid factor (RF) |  | ✓ |  |
| Laboratory study | Lipid Panel |  | ✓ |  |
| Laboratory study | Hemoglobin A1c |  | ✓ |  |
| Laboratory study | Thyroid stimulating hormone (TSH) |  | ✓ |  |
| Laboratory study | Free T4 |  | ✓ |  |
| Laboratory study | 25-hydroxyvitamin D |  | ✓ |  |
| Laboratory study | Serum calcium |  | ✓ |  |
| Laboratory study | EBV anti early antigen IgG, viral capsid IgM, viral capsid IgG, nuclear antigen IgG |  | ✓ |  |
| Laboratory study | D-Dimer |  |  | ✓ |
| Laboratory study | High sensitivity Troponin |  |  | ✓ |
| Laboratory study | High sensitivity C-reactive protein |  |  | ✓ |
| Laboratory study | Procalcitonin |  |  | ✓ |
| Laboratory study | N-terminal pro-brain natriuretic peptide |  |  | ✓ |
| Laboratory study | Insulin C-peptide |  |  | ✓ |
| Laboratory study | Microbiome specimens: sputum, skin swabs, nasal swabs, oral swabs, urine and stool |  |  | ✓ |
